# RGFSAMNet: An interpretable COVID-19 detection and classification by using the deep residual network with global feature fusion and attention mechanism

**DOI:** 10.1101/2024.10.30.24316451

**Authors:** S M Rakib Ul Karim, Diponkor Bala, Rownak Ara Rasul, Sean Goggins

## Abstract

Artificial intelligence has shown considerable promise in fields like medical imaging. Existing testing limitations necessitate reliable approaches for screening COVID-19 and measuring its adverse effects on the lungs. CT scans and chest X-ray images are vital in quantifying and accurately classifying COVID-19 infections. One significant advantage of deep learning models in medical image analysis for detection and classification is that they are a top-notch way to diagnose diseases. For this purpose, we have utilized the power of a deep residual learning network with a global feature fusion technique and attention mechanism to develop our proposed model named “RGFSAMNet” in this study to diagnose the COVID-19 infected patient accurately from a CT scan and chest X-ray images. We have used two publicly available datasets named “SARS-COV-2,” which consists of 2482 CT scan images with two classes, and another chest X-ray dataset that contains 12,576 images with three classes. To check the effectiveness of our model, we have trained and tested the model on two different types of datasets. We also generated the Grad-CAM, LIME, and SHAP visualization based on our proposed model, which can represent the identification of the affected area’s regions in images and describe the model’s interpretability level. These experimental results show that the proposed model architecture can achieve accurate classification of COVID-19 affected CT scans and X-ray images despite a lack of data, with the highest accuracy of 99.60% on test data for CT scans and 99.48% on X-ray image detection and classification. We also developed a web-based User Interface (UI) for the model validation to detect and classify COVID-19 images. Our proposed model exceeds some previous state-of-the-art performance levels. We think our contributions will help clinicians detect and classify COVID-19 images effectively and save human lives.

## 1 Introduction

The phenomenon rapidly disseminated worldwide, resulting in a persistent and intricate public health crisis. The coronavirus releases droplets into the air that are produced by coughing, sneezing, colds, and talking from the nose or mouth of the infected individual, damaging the lungs of others who are nearby. Initially discovered in Wuhan, China, the new coronavirus (COVID-19) started to affect the globe in late 2019 under another name, “SARS-COV-2” Within a month of its first revelation [1] [2]. The virus had gone global and caused a worldwide health emergency. Notwithstanding the most advanced medical therapies, stopping the virus’s spread remained difficult. Primarily by droplets from sneezes and coughing of infected people, COVID-19 damages the respiratory system quickly and could damage the lungs of others who are nearby [3] [4]. It could also cause asphyxia and death from acute oxygen deprivation. As the World Health Organization (WHO) said, the continuous COVID-19 epidemic poses the greatest major threat to healthcare available to mankind. Governments have taken unheard-of steps to stop the virus from spreading. According to published data, as of October 13, 2024, the World Health Organization (WHO) has recorded a total of 776,618,091 confirmed cases of COVID-19, where roughly 7,071,324 deaths have been reported worldwide [5]. Approximately 13.64 billion doses of the COVID-19 vaccine have been given worldwide simultaneously.

Practical diagnostic tools for COVID-19 remain few despite several preventive actions. Usually showing 2-14 days following infection, typical clinical signs include fever, dry cough, tiredness, muscle discomfort, and dyspnea. Still, in rare circumstances, the symptoms could not show up. The fluctuation and spread of the virus challenge proper diagnosis based on symptoms alone, therefore causing possible misinterpretation resulting from symptom overlap with another ailment. Although samples are gathered at current COVID-19 testing facilities, delayed results sometimes impede timely diagnosis and treatment, increasing mortality rates. Reducing mortality rates depends critically on early and accurate COVID-19 diagnosis. Good personal hygiene and social separation lower the infection count, thereby avoiding the overload of hospital systems. Although the significant time and effort involved, laboratory testing, namely reverse transcriptase-polymerase chain reaction (RT-PCR) studies [6], is often regarded as the most precise approach for first diagnosis. These examinations can induce discomfort and provide hazards for both patients and healthcare personnel. Furthermore, these activities demand a substantial time investment, varying from several hours to several days, and are inappropriate for urgent circumstances. Moreover, a significant hindrance is the scarcity of diagnostic tools, especially in impoverished regions.

Additionally utilized to help diagnosis are chest X-rays and CT scans; CT scans show a better sensitivity for identifying lung damage connected to COVID-19 [7]. According to studies, CT scans are helpful diagnostic tools that are rapid and accurate for evaluating lung damage resulting from COVID-19 [8]. To obtain more precise results, Wang et al. [9] implemented a 50-layered ResNet Model on 1136 CT image samples, although they faced trouble distinguishing COVID-19 from normal pneumonia. Artificial intelligence (AI) approaches, including deep learning models, have recently been used to automate COVID-19 identification. These models use medical imaging techniques like chest X-rays and CT scans to forecast the infection’s impact on the respiratory system. They can spot particular abnormalities suggesting COVID-19. When there are few professional radiologists, deep learning can be quite helpful in picture diagnosis and classification. In practical settings, it is an essential instrument for identifying diseases and assessing the degree of lung infections [10] [11].

This study aims to address significant shortcomings in current COVID-19 diagnostic methods by developing a reliable system for detecting COVID-19 from chest X-ray images and CT scans using convolutional neural network (CNN) models. The main objective is to enhance the precision of COVID-19 identification by employing ResNet-50, a Convolutional Neural Network (CNN) structure. The ResNet-50 model was chosen because it can solve the problem of disappearing gradients and better use features already used. Consequently, there is a reduction in the use of parameters and an improvement in the efficiency of the learning process [12]. The dense network technology enables more intricate image processing, gathering visual data on a broader scope in contrast to conventional approaches.

As indicated below, current work provides a substantial contribution to this research in the following contexts:

- Firstly, we employed a novel foster deep residual network with feature fusion and attention model to accurately classify and identify COVID-19 cases. The CT scan and chest X-ray image classification method is a highly sensitive approach for detecting COVID-19, making it a promising option.
- Second, the performance of the suggested model and some baseline pre-trained models has been evaluated using seven distinct metrics, encompassing precision, sensitivity, specificity, accuracy, F1-score, Cohen-Kappa score, and area under the curve. These metrics were employed as part of a comprehensive set of tests.
- Third, this research study addressed a conceptual deficiency by utilizing a larger collection of medical imaging datasets compared to previous research. This is due to the utilization of significantly smaller datasets in previous research.
- Fourth, our proposed method surpasses the existing standard regarding the accuracy and reliability of COVID-19 prediction by using Explainable AI (XAI). This is accomplished by determining the model interpretability capability with the image data.
- Finally, we have developed a web-based User Interface (UI) to validate our proposed model for external images; thus, it would be easy to share our proposed model with end users who can easily and efficiently interact with them.

The remaining parts of the paper are organized as follows: Section 2 focuses on the research already done on COVID-19 detection models. Section 3 provides a comprehensive description of the experimental dataset and a detailed explanation of the methods used. Section 4 outlines the proposed model’s architecture, methods, and experimental setup. Section 5 highlights and analyzes the experimental findings.

Section 6 presents a concise overview and conclusion.

## 2 Related Works

With the emergence of COVID-19, medical image classification has become an issue of concern in the past several years. Researchers immediately shifted their focus to two fronts: the production of vaccinations and improving diagnostic methods to tackle the illness. In recent years, there have been big improvements in radiological image interpretation. This is because doctors need diagnostic tools that are both accurate and efficient. Radiographic imaging, such as Chest X-ray (CXR) and chest computed tomography (CT) scans, has been increasingly crucial in identifying COVID-19 during the pandemic.

CXR images work well for diagnosing lung illnesses, including SARS-COV-2 [13]. In resource-limited settings, it is valuable due to their speed, portability, and cost [14] [15]. CXR can quickly identify typical features of COVID-19, but it faces challenges such as limited sensitivity, radiation exposure, and variability in interpretation due to radiologist experience [16]. In contrast, CT imaging doesn’t involve any invasive processes. It is used to identify lung abnormalities like COVID-19. It also finds early stages of the disease, monitors COVID-19-related anomalies, assesses pulmonary embolism, and guides treatment [17] [18] [19]. CT produces more radiation than CXR. It can be burdensome for its accessibility and cost in certain regions [20]. Several deep convolutional neural network (CNN) models have been suggested to meet this demand. These models use deep learning to assess and categorize medical images accurately, identifying COVID-19 infections using radiographic imaging, making them essential for early diagnosis. Pu et al. [21] and Ozturk et al. [22] employed a 3D-CNN model to detect COVID-19 using CT scan images. Their accuracy rates were 70.20% and 83.90%, respectively. The pandemic has brought attention to the possibilities and difficulties of utilizing deep learning for disease identification. The ongoing research endeavors to overcome data limitations and improve diagnostic capabilities, particularly developing reliable and accurate models for interpreting radiology images. Gülmez et al. [23] designed a new Xception-based neural network optimized with a genetic algorithm (GA) to analyze COVID-19 X-ray images. The GA improves network architecture and settings iteratively. His research outperformed other networks for two, three, and four-class datasets. This study [24] used the ResNet-101 CNN model in deep learning technique, recognizing relevant characteristics in thousands of images during the pre-training phase and then retraining it to detect abnormalities in chest X-ray images. The model only attained an accuracy of 71.9%. Hemdan et al. [25] proposed a deep learning model named COVIDNet to detect the COVID-19 infected patients. He used deep learning algorithms involving end-to-end connected neural networks. The model resulted in an accuracy of 95% on the Covidx dataset, which consists of 127 CXR images from 100 COVID-19 patients. Jain, Gupta, Taneja, and Hemanth et al. [26] applied deep learning techniques to a dataset of 6432 chest X-ray scans from the Kaggle repository, comprising 5467 images for training and 965 for validation. Inception V3, Xception, and ResNeXt models were compared to measure performance using PA or posteroanterior view chest X-rays from both healthy and COVID-19 patients. With 97.97% accuracy, the Xception model did the best. Their research revealed that these models can classify COVID-19, but they do not assure medical precision, and the high accuracy may point to overfitting, requiring further data validation.

COVID-19 disrupts the pulmonary system, resulting in recognizable patchy white lung areas [27]. Some COVID-19 individuals end up with serious breathing problems that require specialist care. Specifically, for those who already have diabetes, hypertension, chronic respiratory ailments, cardiac abnormalities, cancer, or other pre-existing conditions, the SARS-COV-2 virus increases the risk of bad outcomes for those patients [28] [29]. Recent studies have indicated that asymptomatic individuals infected with the coronavirus can still transmit the virus, underscoring the need for effective detection methods [30]. Some deep-learning approaches have been devised to detect pneumonia [31] [32], different categories of thoracic diseases, and other diseases from medical images. Researchers from [33] did some exciting work using chest X-rays and a convolutional neural network (CNN) model called the CDC Net model to find COVID-19, pneumothorax, pneumonia, lung cancer (LC), and tuberculosis (TB). Using three pre-trained CNN models, like ResNet-50, Vgg-19, and Inception v3, they got accuracy rates of 95.61%, 96.15%, and 95.16%, respectively. The confidence-aware anomaly detection (CAAD) model was applied to the X-VIRAL dataset by Zhang et al. [34]. This dataset includes 5,977 participants who tested positive for viral pneumonia and 37,393 subjects who tested negative. According to the study [35], WOCLSA, a multi-layer deep learning model, was presented. This model combines convolutional neural networks (CNN), artificial neural networks (ANN), and long short-term memory (LSTM) networks. The WOCLSA model uses the Whale Optimization Algorithm to find globally optimal solutions. Their models achieved a maximum accuracy of 93% in identifying COVID-19-infected patients. To accurately classify X-ray chest radiographs into numerous or two groups, our study [36] developed a new deep-learning method.

Overall, the suggested model showed an impressive 98.33% accuracy. Several pre-trained transfer learning models have been compared to it after that, including ResNet-101, XceptionNet, DenseNet-121, and VGG-19. Another study [37] introduced a new methodology known as Federated Learning for analyzing X-ray images and got an accuracy of 96.34%. The DBM-ViT (Deep and Broad Model-Vision Transformer) model was proposed by [38], which used depthwise convolutions with different expansion rates to capture global information from Chest X-ray (CXR) and lung CT scan images.

Besides, the ViT module was employed to gather local features for rapid pneumonia diagnosis. The model successfully gathered detection accuracies of 97.25% for CXR and 98.36% for CT images.

Medical diagnostics have increasingly depended on AI-based Internet of Things (IoT) technology. For instance, Yang et al. [39] developed an IoT application that detects Kussmaul breathing in diabetic patients using C-band sensing and a microwave-sensing platform (MSP) to monitor their breathing in real time. It assists physicians in diagnosing diabetic ketoacidosis (DKA). In another study [40], researchers used S-band sensing and Support Vector Machine (SVM) techniques for pattern recognition to track roaming patterns in dementia patients. These advances show how IoT may improve healthcare diagnostics and patient monitoring. The researcher utilized chest CT scans to improve patient outcomes by early identification of COVID-19. To get good results, they used deep convolutional neural networks (CNNs), binary cross-entropy, and transfer learning [41]. The predicted probability for each class is used to compare COVID-19 and normal cases using the binary cross-entropy metric. Optimizers such as Adam and stochastic gradient descent are employed to reduce the impact of overfitting. Singh and Kumar et al. [42] developed a free publicly available deep CNN with ResNet50 for chest radiography infection detection. The suggested model had a 97% accuracy in predicting COVID-19 outcomes.

In the preceding analysis, we examined the written works on both chest X-rays and CT pictures. The study showed that CT scans better detect COVID-19 than chest X-rays. Due to its adaptability, the using a deep residual network with global feature fusion and an attention mechanism-based model beats all other techniques. We have proposed a model named “RGFSAMNet” to increase COVID-19 class recognition accuracy.

## 3 Materials & Methods

### 3.1 Data Collection

SARS-COV-2 [43] and the Chest radiography [44] [45] datasets were publicly available datasets that we employed to evaluate our proposed model CONet to detect the presence of COVID-19. The SARS-COV-2 dataset contains CT scan pictures of COVID-19 patients and healthy persons. Hospitalized patients in Sao Paulo provided these statistics. The images depict persons who have been hospitalized due to SARS-COV-2 infection (COVID) and those who have other lungs damaged by the virus, including 60 women and 60 men with abnormalities. It was gathered from a total of 120 patients who participated. This dataset comprises 2482 CT scans, including 1252 from COVID-19-positive patients and 1230 from COVID-19-negative patients.

The suggested model CONet detects COVID-19 using chest radiography and SARS-COV-2. A chest radiography dataset includes COVID-19, pneumonia, and healthy patient X-rays. Pneumonia, COVID-19, and Normal each have 4192 images, totaling 12,576. We need these images to train and evaluate our system for identifying COVID-19 from other lung illnesses. Image samples from multiple situations allow a robust evaluation of the model’s generalization and performance on unseen data.

Both datasets aim to support scientific research on artificial intelligence systems capable of analyzing CT scans and chest radiography pictures to detect SARS-CoV-2 infection in individuals.

### 3.2 Methodology

The proposed methodology consists of some main steps, including data preprocessing, model development, hyperparameters setting, and training and testing the proposed models on the dataset. Firstly, we load the lung CT scan and Chest X-ray image datasets into a working environment. Once the dataset is loaded, the data pre-processing operations, such as image resizing, data normalization, and data augmentations, are performed. Then, we utilized the deep residual learning context with global feature fusion concepts and attention mechanism to build the proposed deep learning model named “RGFSAMNet”. Furthermore, we will be dividing the dataset into training and testing. In this model, we divide the dataset into 80% for the training set, 20% from the training for validation, and the remaining 20% for testing. We will develop a tailored approach to train our new model in batches and allow it to run for a few epochs. Ultimately, it is necessary to assess the model’s performance by evaluating it using the testing dataset. The semantic representation of the proposed methodology is shown in Figure 2:

**Fig 1.**
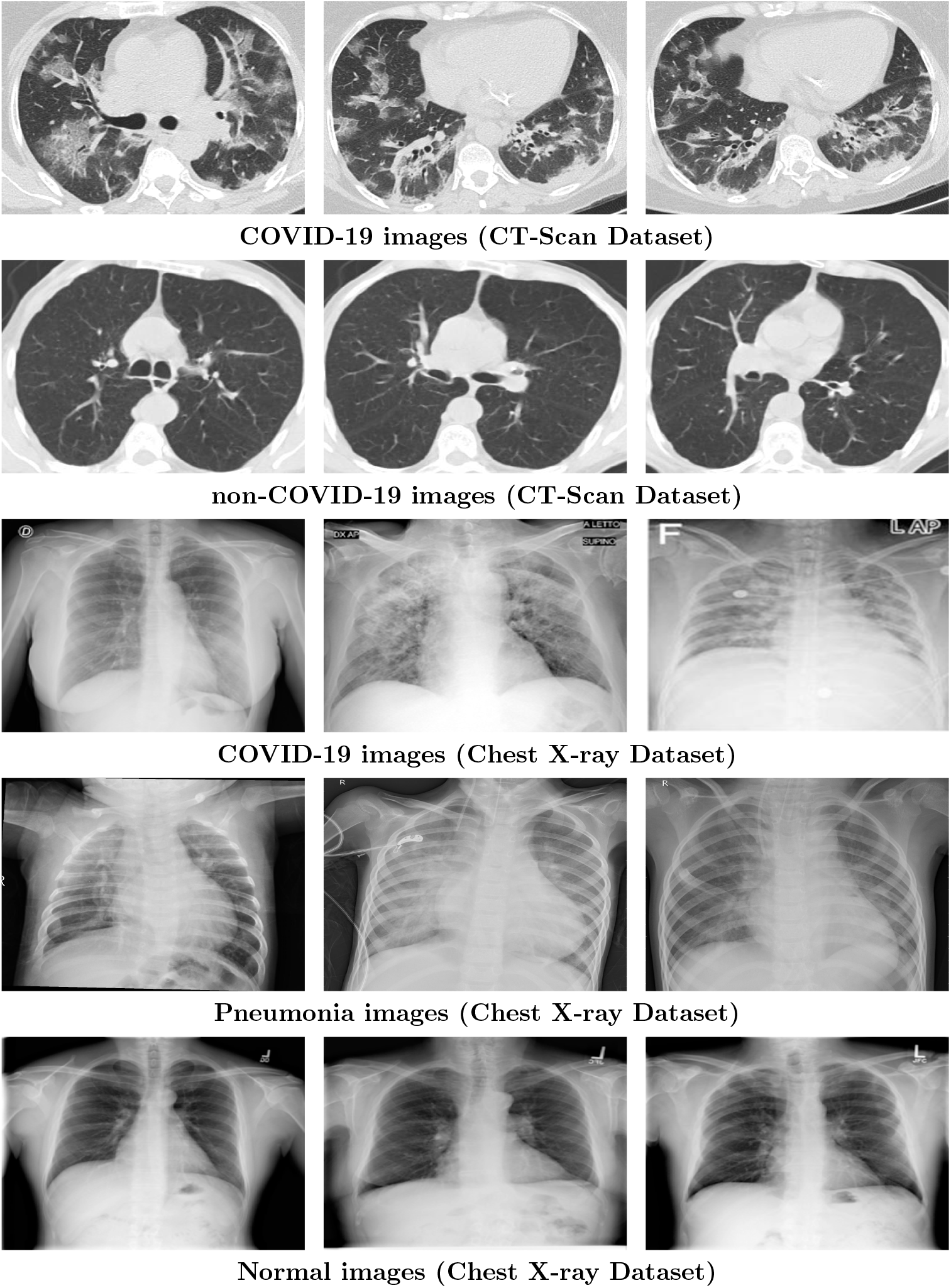
Sample images for both datasets

**Fig 2.**
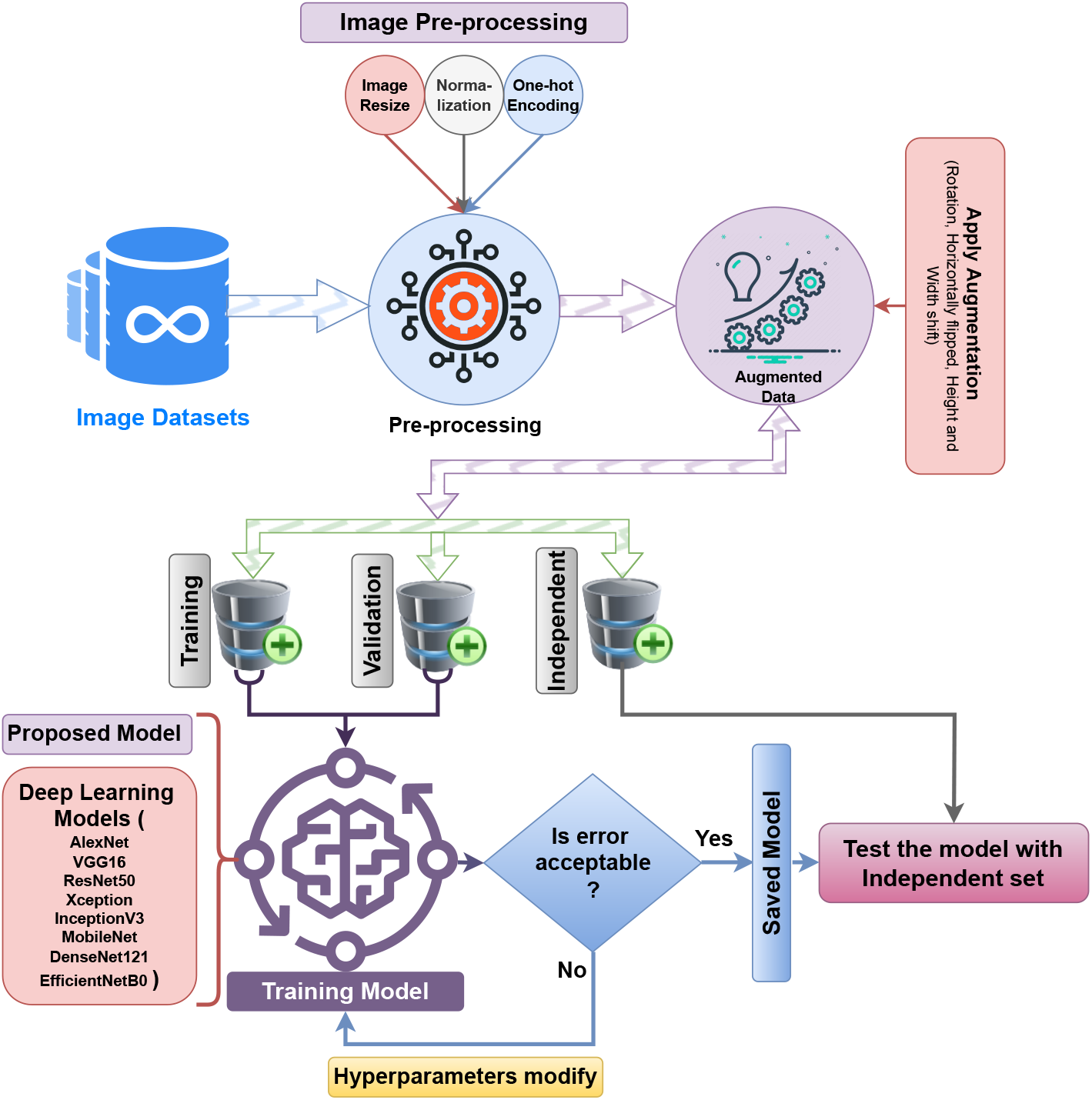
Schematic representation of the proposed methodological framework.

#### 3.2.1 Data Pre-processing

Data preprocessing involves several processes, including extracting labels from images, converting images to RGB format, conducting feature scaling, and resizing images.

##### Feature scaling

Feature scaling usually applies to the independent variables of input data to standardize them within a fixed range such as [0, 1]. In this method, the CT scan and X-ray pictures in the dataset are initially converted to RGB format and then enlarged to 224x224 pixels using OpenCV procedures. The conventional range of image pixel values is [0, 255]. Here 255 is the maximum value. At the beginning, two distinct datasets are loaded with images and records. Subsequently, the data is transformed into NumPy arrays to facilitate feature scaling. Max normalization changes the pictures’ data to the range [0, 1] by dividing each pixel value by 255. On the other hand, the label data is encoded one-hot to change the category labels into binary format [46]. As a result, the normalized number is shown by the letter *x*^*′*^.

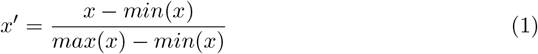

where *x* is the original intensity of the image.

##### Data splitting

We used a local directory to access the dataset and directly accessed them using the “operating system” library of Python and subsequently processed with the OpenCV library. The images were then converted into appropriate data types to facilitate model training. It’s necessary to partition the dataset into training and testing subsets. For this model, the dataset is divided into 80% for the training set, 20% from the training set for validation, and 20% testing set [47].

##### Data Augmentation

In data augmentation, we tried to expand our dataset to enhance its diversity artificially. The primary goal is to introduce variations through minor modifications of the original data. An ImageDataGenerator class is provided by the Keras deep learning framework to fit the model through image data augmentation. In this study, we applied data augmentation to the training data using the ImageDataGenerator class, generating multiple versions of the same images. This approach is particularly advantageous when working with a limited dataset [48]. The augmentation was only applied to the training data as detailed in Table 1, as well as the visualization of the augmented samples, is shown in Figure 3 and here we have chosen to:

**Table 1.** Data augmentation parameters that we utilized in this research.

| Data-augmentation parameters name | Parameters value | Action |
| --- | --- | --- |
| Rotation range | 20 | The input data are produced by rotating from -45 to 45 degrees. |
| Horizontal flip | True | Randomly flip inputs horizontally. |
| Height shift range | 0.2 | Randomly shifted in height by 20%. |
| Width shift range | 0.2 | Randomly shifted in width by 20%. |

**Fig 3.**
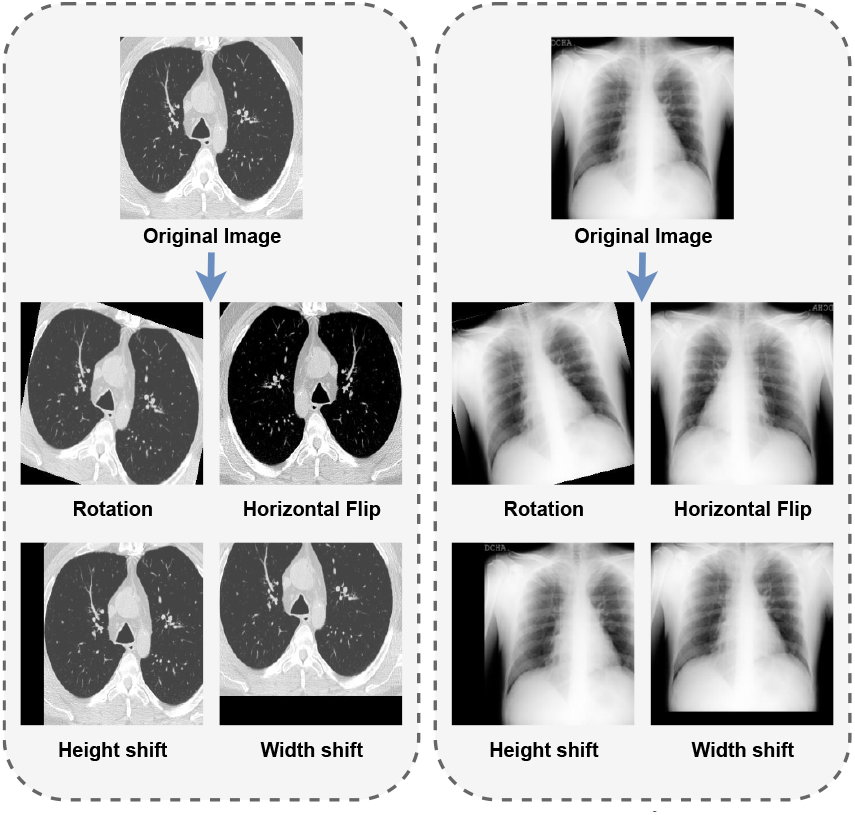
Samples of augmented images of Chest CT-scan & X-ray.

#### 3.2.1 Deep Learning Models

Recent advances in deep learning processes have transformed artificial intelligence. “Deep states” refers to the model’s network layer sizes. The utilized pre-trained deep learning models are given below:

##### AlexNet

AlexNet is a pivotal CNN model that significantly advanced deep learning. It has five convolutional layers and three fully connected layers, using ReLU activation and max-pooling. Each convolutional layer is defined by *f* (*x*) = max(0, *W*∗*x* + *b*), where *x* is the input, *W* is the weight matrix, *b* is the bias, and ∗denotes convolution [49]. The network’s architecture includes 96 11x11 filters with a 4x4 stride, max pooling with a 3x3 window and 2x2 stride, followed by layers with 256 5x5, 384 3x3, 384 3x3, and 256 3x3 filters, respectively. After flattening, three fully connected layers with 4096, 4096, and 2 neurons follow, with a softmax activation for classification. Dropout layers with a 0.5 rate are used after the first two fully connected layers to prevent overfitting. Max-pooling is used to reduce the spatial dimension of the representation to decrease the number of parameters and calculations in the network.

##### VGG16

VGG16 is part of CNN with 3 fully connected layers, including 13 convolutional and 16 layers. It uses 2× 2 max pooling with a stride of 2 and 3× 3 convolutional filters. The input size is fixed at 224 ×224 × 3 for RGB images [50]. The model is initialized with pre-trained weights from the ImageNet dataset, excluding the top fully connected layers. The output is flattened and passed through two custom dense layers: the first with 128 units and ReLU activation, and the second serving as the output layer with 2 units and softmax activation for binary classification (COVID-19 positive or negative) based on X-ray or CT scan images. All layers of the base model are frozen (layer. trainable = False), allowing the model to leverage features learned from ImageNet.

Mathematically, the output of each convolutional layer is given by

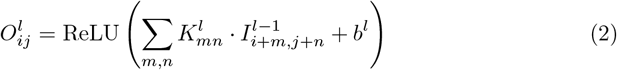

where 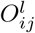 is the output at location (*i, j*) in layer 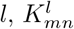 is the kernel weight at location (*m, n*) in layer *l*, 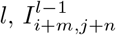 is the input from the previous layer at location (*i* + *m, j* + *n*), and *b*^*l*^ is the bias term for layer *l*. The ReLU activation function is applied element-wise. The fully connected layers are represented as

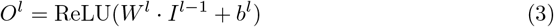

where *W*^*l*^ is the weight matrix, *I*^*l*−1^ is the input from the previous layer, and *b*^*l*^ is the bias term.

##### ResNet-50

ResNet-50 is a deep neural network with 50 layers designed for efficient training. It utilizes residual blocks to address the vanishing gradient problem, with the key equation being *H*(*x*) = *F* (*x*) + *x*, where *H*(*x*) is the desired mapping, *F* (*x*) is the residual mapping, and *x* is the input. Shortcut connections help in gradient propagation, enabling the training of deeper networks *j* and *K* is the number of classes [12]. For COVID-19 detection, a softmax function is used in the final layer to output probabilities for positive or negative classes, with the equation 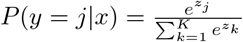, where *z*_*j*_ is the logit for class.

##### Xception

The Xception model, a deep learning architecture, employs depthwise separable convolutions, a technique from the Inception architecture [51]. It splits the convolution operation into depthwise and pointwise convolutions. The depthwise convolution applies a filter to each input channel, while the pointwise convolution uses 1x1 convolutions to combine the depthwise convolution results. The mathematical representation is:

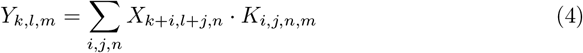

where *X* is the input tensor, *K* is the kernel tensor, and *Y* is the output tensor. The indices *k, l* represent spatial dimensions, *n* is the channel dimension, and *m* is the output channel dimension.

The Xception model is initialized with pre-trained ImageNet weights and configured without the top layer for transfer learning. A flattened layer transforms the 3D output to a 1D tensor, followed by a Dense layer with 128 units and ReLU activation for feature learning. A final Dense output layer with two units and softmax activation is used for binary classification, such as detecting COVID-19. The base model’s layers are frozen to prevent weight changes during training, allowing only the added layers to learn new weights.

##### MobileNet

MobileNet is a classification of efficient convolutional neural networks (CNNs) specifically created for mobile and integrated vision tasks. It employs depthwise separable convolutions to reduce the number of parameters and computational costs while maintaining accuracy. The architecture is controlled by two hyperparameters: the width multiplier (*α*) and the resolution multiplier (*ρ*), which adjust the network’s depth and input resolution, respectively [52]. In a depthwise separable convolution, a depthwise convolution applies a separate filter to each input channel, followed by a pointwise convolution that combines the channels. This reduces the computational cost from *O*(*HWDK*^2^*M* ) for a standard convolution to *O*(*HWDK*^2^ + *H*^*′*^*W*^*′*^*DM* ) for a depthwise separable convolution.

Initializing MobileNet with pre-trained ImageNet weights and ignoring the top classification layer, COVID-19 is detected from X-ray or CT scan pictures. The base model has frozen all layers; the input shape is 224× 224× 3. Personalized layers are applied on top, comprising a Flatten layer, a dense layer with 128 neurons and ReLU activation, and a last dense layer with 2 neurons and softmax activation for binary classification. After that, the model is assembled and trained using the CT or X-ray image dataset.

##### InceptionV3

InceptionV3 is a convolutional neural network (CNN) architecture optimized for efficient image recognition tasks. It utilizes “Inception modules,” which allow the network to learn from different scales of the input image simultaneously. Each Inception module contains convolutional layers with varying kernel sizes (e.g., 1x1, 3x3, 5x5) and a pooling layer concatenated together. This design enables the network to capture both local and global features in the image [53].

Mathematically, the output of a convolutional layer in an Inception module is expressed as:

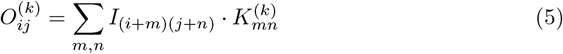

where 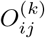 is the output of the *k*-th filter at position (*i, j*), *I* is the input image, and *K*^(*k*)^ is the kernel of the *k*-th filter.

InceptionV3 can be a feature extractor for COVID-19 detection using X-ray or CT scan images. It can learn complex patterns indicating the presence of the virus. The extracted features are further processed through additional layers (Dense, BatchNormalization, Dropout) to classify the images as COVID-19 positive or negative. The final Dense layer with softmax activation produces the probabilities for each class.

##### DenseNet121

DenseNet121 (Dense Convolutional Network with 121 layers) is a deep neural network architecture with feed-forward connections between each layer and all other levels. This design results in a highly dense network, hence the name DenseNet. Every layer in DenseNet takes as inputs the feature maps from every other layer, and every layer after that uses its feature maps as inputs [54].

The output of the *i*-th layer in a DenseNet, denoted as *x*_*i*_, can be expressed as:

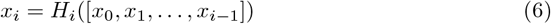

The concatenation of the feature maps from all previous layers is represented by [*x*_0_, *x*_1_, … , *x*_*i*−1_], and *H*_*i*_ is a function that is made up of processes like Pooling, Convolution, Batch Normalization (BN), and Rectified Linear Unit (ReLU) activation.

DenseNet121 can be used as a feature extractor for COVID-19 detection from X-ray or CT scan images. Pre-trained on ImageNet, the model helps capture generic features from medical images. The code provided initializes a DenseNet121 model with pre-trained weights and modifies it for binary classification, distinguishing between COVID-19 positive and negative cases. Adding a flattened layer, a Dense layer with ReLU activation, and a last Dense layer with softmax activation for classification helps the model to be customized for this particular task.

##### EfficientNetB0

EfficientNetB0 is a convolutional neural network architecture from the EfficientNet family, known for its efficiency in accuracy and resource usage [55]. It scales network dimensions using a compound coefficient, balancing depth, width, and resolution. The formula for scaling is *d* = *α*^*ϕ*^, *w* = *β*^*ϕ*^, *r* = *γ*^*ϕ*^, with *ϕ* controlling overall scaling, and *α, β*, and *γ* as constants determined through grid search. In our application, EfficientNetB0, with pre-trained ImageNet weights, serves as the base model. It’s followed by a Flatten layer, BatchNormalization and Dropout layers for regularization, and Dense layers with ReLU and softmax activations for binary classification in COVID-19 detection from X-ray or CT scan images.

#### 3.2.3 Proposed Model Descriptions

The latest developments in deep learning systems have impacted artificial intelligence by making the models more sophisticated and accurate in a wide range of tasks. In a neural network, Deep states are a term that measures the number and size of the layers in the network, indicating its depth. The models may learn the features at a high level by using this amount of depth, which can improve their performance on complex tasks.

The entire structure is represented by a deep residual learning mechanism-based block (ResNet-50) as the base model. Deep CNN model mainly comprises three main layers: Convolution, Max-pooling, and Dense. For extracting features, the convolution layer uses filters from the input data. The max-pooling layer usually increases computational performance by minimizing the size of these features. The dense layer connects all previous layers. The dense layer is also known as a fully connected layer.

The deep CNN models can be optimized for classification and detection by modifying hyperparameters.

The proposed RGFSAMNet model architecture is shown in Figure 4 and described below in detail.

**Fig 4.**
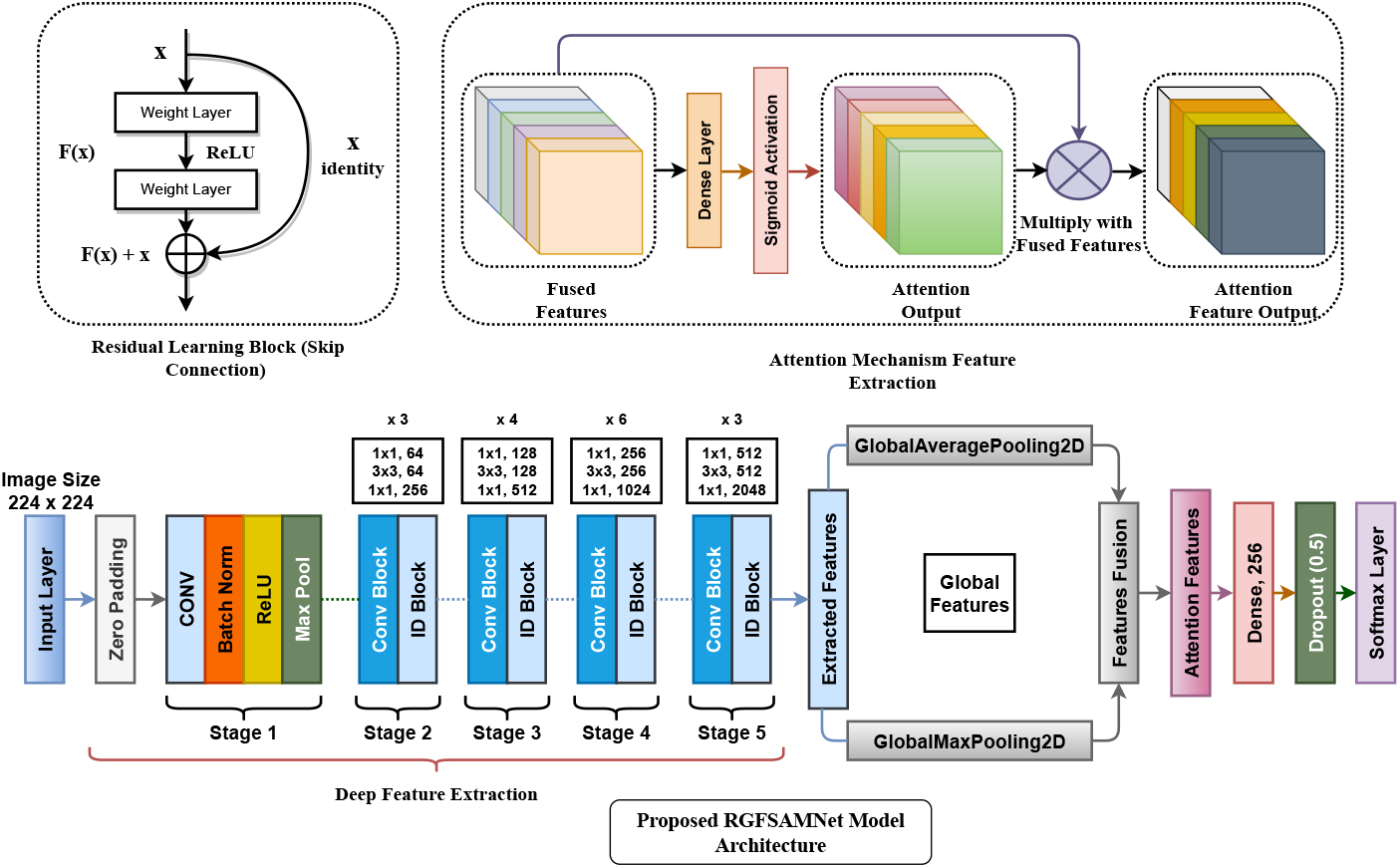
Schematic architecture of the proposed model.

##### Input Layer

The input layer of the CNN contains the numerical values representing the pixels of the input image. The input is a 224x224x3 pixel array for a 224x224 dimension image with three color channels, specifically the RGB color channel [56].

##### Convolutional Layer

The Convolutional Layer is the key component of CNN. The Convolutional Layer produces a new image that displays the features of the input image. The convolutional layer applies a filter to each input image during the process. The filter on this layer is a 2-dimensional array with dimensions of 5x5, 3x3, or 1x1.

Then the filter generates a feature map, later used by the Action Layer [57]. The 2D convolutional operator for the received image (X) and kernel (k) is:

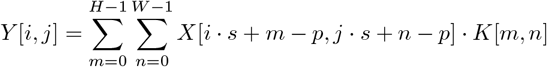

Where *Y* [*i, j*] is the output at position (*i, j*). *X*[*i· s* + *m*− *p, j· s* + *n*− *p*] is the input at position (*i· s* + *m* −*p, j ·s* + *n* −*p*), *K*[*m, n*] is the kernel at (*m, n*), *H* and *W* are the height and width of the kernel, *i* and *j* are the output coordinates, *s* is the stride (defining the step size of the kernel as it moves across the input image) and *p* is the padding (the number of pixels added to the border of the input image).

##### Base Model

For our proposed model, we used the ResNet-50 pre-trained model as the base model. ResNet-50, one of the major advancements in deep learning, is used in this project. Training the model before ResNet came into existence seemed to be one of the hardest tasks due to fading gradients. Skip connection is one of the major strengths of the ResNet model. ResNet-50 has 50 neural network layers. The network takes the input image in multiples of 32 as width and height. ResNet-50 is a modified version of the Residual Network (ResNet) structure, specifically developed to facilitate the training of extremely deep neural networks. The network is composed of 50 layers, which include convolutional, batch normalization, and ReLU layers. Additionally, shortcut connections enable the network to bypass one or more levels. The fundamental mathematical principle behind ResNet is the incorporation of residual blocks, in which the input is combined with the output. This approach effectively addresses the issue of the vanishing gradient problem and facilitates the training of more complex networks [12].

ResNet-50, like other ResNet architectures, is built on the concept of residual learning. The left upper part of Figure 4 shows the block diagram of the residual learning block. The core idea is expressed mathematically as:

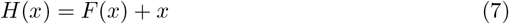

Here, *H*(*x*) is the desired underlying mapping that the network aims to learn, *x* is the input to a residual block, and *F* (*x*) is the residual mapping that the network needs to learn. The term *F* (*x*) + *x* represents the output of the residual block, where *F* (*x*) is the output of the stacked non-linear layers (e.g., convolutional, batch normalization, and ReLU layers) and *x* is the shortcut connection that bypasses these layers. In a network with multiple layers like ResNet-50, this approach is repeatedly used in many residual blocks, allowing the network to learn increasingly complex mappings while mitigating the problem of vanishing gradient. Shortcut connections aid in the propagation of gradients during backpropagation, hence streamlining the training of deeper neural networks.

##### Global Feature Fusion

Global feature fusion is generally accepted to combine multiple global pooling strategies, such as Global Average Pooling (GAP) and Global Max Pooling (GMP), for extracting more comprehensive and robust features from a deep CNN model. These techniques will be able to retain both the global spatial information and most significant feature activations [58].

Global Average Pooling (GAP) [59] is a pooling operation that calculates the average of each feature map across the spatial dimensions, reducing each feature map to a single scalar value. This operation effectively compresses the spatial dimensions, maintaining the global structure of the feature maps. For a given feature map *F*∈ℝ^*H*×*W*×*C*^ with height *H*, width *W* , and *C* channels, the GAP for each channel *c* can be computed as:

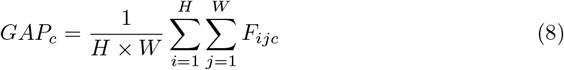

where *F*_*ijc*_ is the value of the feature map at position (*i, j*) in channel *c*. The result *GAP*_*c*_ is a scalar value representing the average of the entire feature map for the *c*-th channel. This operation reduces the feature map *F*∈ℝ^*H*×*W*×*C*^ to a vector *GAP* ∈ℝ^*C*^ .

Global Max Pooling (GMP) [60] selects the maximum value from each feature map across the spatial dimensions. This helps in capturing the most prominent or activated feature in each channel, making it useful for identifying strong activations.

For a feature map *F* ∈ℝ^*H*×*W*×*C*^ , the Global Max Pooling (GMP) for each channel *c* is computed as:

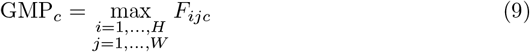

where GMP_*c*_ is the maximum value from the entire feature map for the *c*-th channel.

This operation reduces the feature map *F*∈ ℝ^*H*×*W*×*C*^ to a vector GMP∈ ℝ^*C*^ .

The idea of global feature fusion is to combine the features extracted by both GAP and GMP, leveraging the benefits of both approaches. While GAP captures the overall distribution of the feature map, GMP focuses on the most salient features. One simple fusion method is concatenation, where the outputs of GAP and GMP are concatenated along the channel dimension:

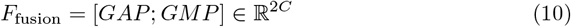

Here *GAP* ∈ℝ^*C*^ is the vector resulting from global average pooling. *GMP*∈ℝ^*C*^ is the vector resulting from global max pooling. *F*_fusion_ ∈ℝ^2*C*^ is the concatenated feature vector, containing both average and max-pooled features.

##### Attention Mechanism

The attention mechanisms in image classification give the model a way to learn which parts of the input are more important for the task at hand; this is therefore helpful for performance by effectively underlining features that will be crucial. A short description of attention is that it calculates the weighted sum of features in a manner such that more important features have higher weights. In an image, the features are the pixel values in feature maps that are created by a deep learning model [61].

The attention function can be summarized as:

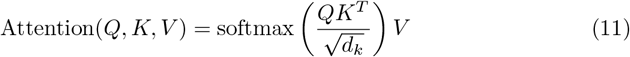

Where 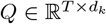 is the query matrix. 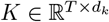 is the key matrix. 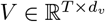 is the value matrix. *d*_*k*_ is the dimensionality of the key vectors. The softmax function ensures that the attention weights sum to 1.

##### Activation Layer

The layers are reduced into 512 and a pyramid-like shape. The major role of this layer is the reduction and the activation function used here is the ReLU. The activation function is responsible in a neural network for converting the total weighted input from the node into the node or output activation for this input [62]. The equation of the ReLU function is represented as:

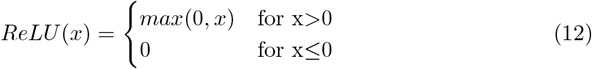

where *x* is the neuron’s input value.

##### Droupout Layer

One of the techniques for neural network regularization is known as dropout. It prevents overfitting due to a section of neurons being randomly ‘dropped out’ from the forward pass during training. This dropout is made with some probability and is applied independently to every neuron in a layer. In our proposed model, we used a dropout rate of 0.3, which means 30% of randomly selected neurons were disabled during model training [63].

##### Dense Layer

The Dense Layer is one of the most important neural network architectures, providing flexibility and expressiveness to learn from the data. This ability makes all neurons in this layer connect to the previous layer, hence helping to model complex relationships and thus quite indispensable in deep learning. In a dense layer, each neuron is connected to every input feature, which is what makes it highly expressive, hence able to learn complex patterns [64].

##### Output Layer

The output layer here is named as ‘Prediction’. The output of the dense layer is fed as input and the output is obtained as a binary value of either masked or non-masked. The output of this layer is fed directly as output to the model [65]. The softmax function is defined as:

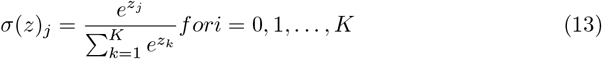

This function takes a *K*-dimensional input vector *z* and returns a *K*-dimensional vector of values within the range 0 to 1 that sum to 1.

### 3.3 Explainable AI

By making machine learning models more transparent and interpretable, XAI (Explainable Artificial Intelligence) methods like Grad-CAM, LIME, and SHAP can be vital in healthcare for COVID-19 detection. Building trust in AI-based diagnostic systems is essential, and these techniques help researchers and clinicians understand why a particular prediction was made. The XAI methods can help identify COVID-19 in medical images and patient data by creating graphical representations of model decisions. Along with facilitating a more precise diagnosis, this also aids in the discovery of possible disease-related biomarkers or patterns. Ensuring patient safety, minimizing errors, and fostering collaboration between AI systems and healthcare professionals are crucial in a field as critical as healthcare, where decisions directly impact patient outcomes. Having explainable AI is a must.

#### 3.3.1 Grad-CAM as XAI

Grad-CAM is also a powerful technique for visualizing and understanding decisions of CNN-based models. In fact, by highlighting the important regions of an image, Grad-CAM shows how a model reaches a particular decision; therefore, it is one of the most important methods in model interpretability, debugging, and trustworthiness.

Let *A*^*k*^ denote the *k*-th feature map (of size *H*× *W* ) of the final convolutional layer, and let *y*^*c*^ represent the score for the class *c*. We first compute the partial derivatives of the class score *y*^*c*^ with respect to the feature maps *A*^*k*^:

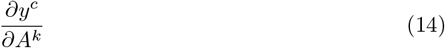

These gradients indicate how much the output for the class *c* changes with respect to the activation in each feature map.

Next, we calculate the importance (weight) of each feature map by performing global average pooling over the spatial dimensions (height and width) of the gradients:

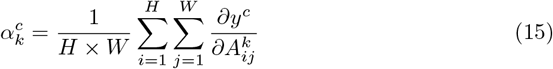

Here, 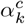 represents the importance of feature map *k* for predicting class *c*.

Then the class activation map is computed by taking a weighted sum of the feature maps, where each feature map *A*^*k*^ is multiplied by its corresponding weight 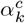:

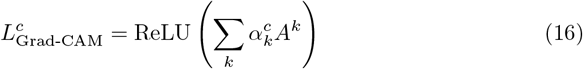

The ReLU activation is applied to keep only the positive contributions, as negative values are not considered relevant to the class prediction.

#### 3.3.2 LIME as XAI

The healthcare industry can significantly benefit from LIME’s (Local Interpretable Model-agnostic Explanations) [66] ability to detect COVID-19 by offering interpretable insights into the predictions made by machine learning models. Our proposed model uses LIME to describe the COVID-19 detection and prediction of a model trained on medical images, possibly X-rays or CT scans. By modifying the “LimeImageExplainer” class, we can justify picture classifications, detailing which parts of the picture had the most significant impact on the model’s final verdict.

By better understanding why the model assigned a specific image a positive or negative COVID-19 classification, clinicians can make better-informed decisions regarding patient diagnosis and treatment planning. Because of LIME’s openness, healthcare providers can trust and validate AI-based predictions, which improves patient care.

Here the LIME as XAI is represented as follows:

- *f* is the model to be explained
- *x* is the input instance to be explained
- *π* is a proximity measure between instances
- *L* is an interpretable model
- 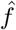 is the locally interpretable model (LIME model)
- Ω(*L*) is the complexity of the interpretable model

The explanation is obtained by solving the following optimization problem:

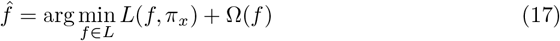

Where *L*(*f, π*_*x*_) is a loss function that measures the difference between the predictions of the interpretable model and the original model for the instance *x*, Ω(*f* ) is a regularization term that penalizes complex models.

#### 3.3.3 SHAP as XAI

By illuminating the model’s reasoning behind medical image classification decisions, SHAP (SHapley Additive exPlanations) [67] can greatly help the healthcare industry’s search for COVID-19. We use SHAP to explain the model’s predictions on a picture of possible COVID-19 symptoms. The SHAP values help medical professionals see which parts of the picture matter for diagnosis because they show how much each pixel contributed to the final product.

For AI-based diagnostic systems to gain confidence from users, their decisions must be interpretable so that doctors can validate them using their knowledge. In addition, SHAP can aid in the identification of particular radiological patterns or characteristics indicative of COVID-19, allowing for faster and more precise diagnoses. SHAP’s interpretability is essential to guarantee patient safety and improve the efficacy of diagnostic procedures in the fast-paced and high-stakes healthcare environment.

- Let *f* : ℝ^*d*^ →ℝ be the function representing the model’s prediction, where *d* is the number of features. The SHAP value for feature *i* in the context of a specific prediction *x* is given by:

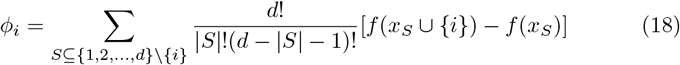

Where *x*_*S*_ represents a subset of features excluding feature *i*, The sum is over all subsets *S* of features excluding *i*, |*S*| denotes the number of features in subset *S, f* (*x*_*S*_ ∪ *{i}*) is the model’s prediction when including feature *i* in the subset *S, f* (*x*_*S*_) is the model’s prediction without including feature *i* in the subset *S*.

### 3.4 Proposed Model Training

The features extracted from the image are provided as input to the convolutional neural network layers. Here, we use the ResNet-50 deep convolutional neural network as our base model, and this network aids model training with its 50 levels of depth. The base model is not trained with any predefined weights. The shape of the input is defined as the image size of the processed image, which is 224*224 pixels. The base model’s output is put through global average pooling. The ReLU activation function is used as the model’s input, and the Softmax function is used as the output. The model is then optimized with the Adam optimizer with a learning rate of 0.003; the loss here is considered to be the categorical cross-entropy, and the evaluation metrics of every training epoch are calculated with accuracy. The model is then trained with the train and validation dataset with a batch size of 32, and the epochs considered here are 150 and 100.

#### 3.4.1 Utilized model parameters

The hyperparameters in machine learning control the learning process. The proposed model and all the deep learning models used had numerous parameters, leading to many possible architectural variations. Additionally, training these models with a large dataset was time-consuming. Given our limited computing resources, hyperparameter optimization would have been impractical. Therefore, we used common hyperparameter values and explored other techniques to enhance model evaluation.

The set of hyperparameters used in the proposed model were: i) optimizer (ii) loss function, ii) the number of epochs n, iii) batch size b, and v) learning rate. The following explains the reasoning behind the hyperparameter values selected:

##### Optimizer

Optimizers determine how to update model parameters to minimize loss function during machine learning training. Adam [68], SGD [69], and RMSProp are popular optimizers. Adam employs the estimation of the first and second moments of the gradients to dynamically modify the learning rates for each parameter using RMSProp and momentum optimization. RMSProp employs a rolling average of squared gradients to adapt the learning rates of individual parameters according to the magnitude of the gradient. Stochastic Gradient Descent (SGD) optimizes the model parameters by utilizing the gradient of the loss function concerning the weights, which are calculated on a small subset of training data. This approach can accelerate the convergence process, especially with extensive datasets. The selection is contingent upon the problem’s nature, the dataset’s magnitude, and the available computational resources. Every optimizer possesses distinct benefits and is well-suited for certain situations.

- **Adaptive Moment Estimation (Adam)** is the predominant optimization algorithm currently employed for training deep neural networks due to its simplicity in implementation, high computational efficiency, and exceptional performance in handling extensive data and parameters. Adam can be thought of as a stochastic gradient descent with momentum combined with RMSprop. For this investigation, we employed the Adam optimizer with *β* values of 0.9 and 0.999, *ϵ* of 0.1, and weight decay of 0.01. The mathematical formula for the Adam optimizer update rule is as follows: **Initialize parameters**
  – *θ*: Model parameters to be optimized.
  – *α*: Learning rate.
  – *β*_1_: The exponential decay rate for the initial instant estimate is determined (typically set to 0.9).
  – *β*_2_: The rate of deterioration for the second moment estimate follows an exponential pattern (typically set to 0.999).
  – *ϵ*: Small constant to prevent division by zero (typically set to 10^−8^).
  **Initialize moment variables:**
  – *m*_0_ = 0 (Initial first moment vector).
  – *v*_0_ = 0 (Initial second moment vector).
  **For each iteration** *t* = 1, 2, …:
  1. Compute gradient *g*_*t*_ on the current mini-batch.
  2. Update biased first moment estimate:

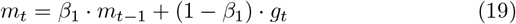
  3. Update biased second raw moment estimate:

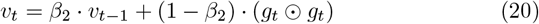

(Here, ⊙ denotes element-wise multiplication)
  4. Correct bias in the first moment:

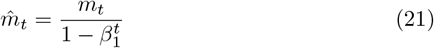
  5. Correct bias in the second raw moment:

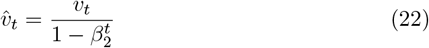
  6. Update parameters:

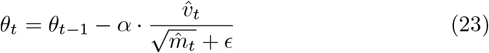
- **Root Mean Square Propagation (RMSProp)** is an optimization strategy employed to adjust the weights of a neural network according to the gradient of the loss function. The approach is an adaptive learning rate technique that modifies the learning rate of each parameter by considering the average of the squared gradients for that parameter across time. This helps to overcome the problem of vanishing or exploding gradients and can lead to faster convergence. The update rule for RMSProp can be represented mathematically as follows:

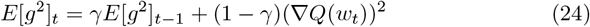

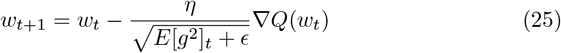

where *w*_*t*_ is the current set of weights of the model at iteration *t, η* is the learning rate, ∇*Q*(*w*_*t*_) represents the derivative of the loss function with respect to the weights, *E*[*g*^2^]_*t*_ represents the average of the squared gradients, which fade exponentially, *γ* is the decay rate (typically set to 0.9), *ϵ* is a minute amount that is added to the denominator in order to avoid division by zero.
- **Stochastic Gradient Descent (SGD)** is a modified version of the gradient descent optimization technique that is frequently employed to train machine learning models, particularly in the field of deep learning. It works by updating the model weights iteratively based on the gradient of the loss function concerning the weights, computed on a small subset of the training data (mini-batch), rather than the entire dataset. This approach can lead to faster convergence than standard gradient descent, especially for large datasets. The update rule for SGD can be represented as follows:

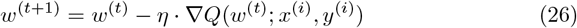

where *w*^(*t*)^ is the current set of weights of the model at iteration *t, η* represents the learning rate, controls the size of the update step, ∇*Q*(*w*^(*t*)^; *x*^(*i*)^, *y*^(*i*)^) is the gradient of the loss function *Q* concerning the weights, computed on a single example (*x*^(*i*)^, *y*^(*i*)^) from the training dataset. **Loss function** A loss function called categorical cross-entropy is employed in multi-label categorization. This is when only one category is applicable for each data point. This function performed flawlessly in this scenario, as each case could only be classified under either the two or three categories. [71].

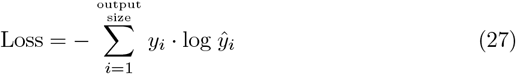 Where ŷ_*i*_ and *y*_*i*_ represent the *i*_*th*_ scalar and its corresponding target value, respectively. The term “output size” denotes the number of scalar values the model generates. **Epochs** Upon conducting numerous preliminary trials with values of 50, 100, 150, and 200, the most optimal results were achieved with 150 and 100 epochs. **Batch Size** A batch size of 32 yielded the most optimal results after several initial tries with values of 32, 64, and 128. **Learning Rate** An optimizer with a learning rate annealing strategy was used. Implementing a progressively declining learning rate during the training process facilitated the efficient attainment of the global minimum of a loss function [72]. The initial learning rate was established at 0.003 and was diminished by a factor of 0.75 if there was no enhancement in validation accuracy after ten epochs, as specified by the patience parameter.

### 3.5 Environment Setup

The software was implemented using Python3. The GPU was selected over the CPU for its superior speed and efficiency in training deep-learning models. The experiments were performed on an Apple Macbook Pro utilizing an Apple silicon M2 Max GPU.

### 3.6 Evaluation metrics

We evaluated the performance of our proposed model using a range of metrics, including precision, accuracy, F1 score, recall/sensitivity, specificity, Cohen’s kappa score, and Area Under the Curve (AUC). The confusion matrix, which includes the values for True Positive (TP), True Negative (TN), False Positive (FP), and False Negative (FN), was utilized to derive these metrics [73]. By applying these metrics, we comprehensively evaluated the model’s ability to detect instances accurately. The model’s efficiency was determined using the following formula:

- **Precision:** It quantifies the precision of the favorable forecasts. It can be characterized by-

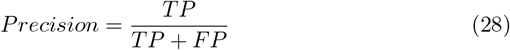
- **Sensitivity/Recall:** It determines the proportion of true positives correctly identified. It can be characterized by-

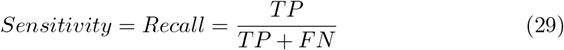
- **Specificity:** It determines the proportion of true negatives correctly identified. It can be characterized by-

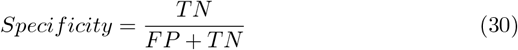
- **Accuracy:** Accuracy is a metric that quantifies the extent to which a model is right. It can be characterized by-

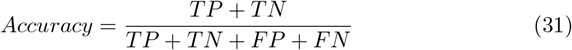
- **F1 Score:** This measure is the reciprocal of the arithmetic mean of precision and recall. It can be characterized by-

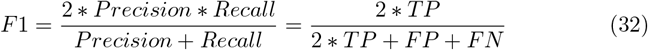
- **Cohen-Kappa Score:**Cohen’s Kappa Score [74] enumerates the level of agreement between two raters or classifiers, using a scale ranging from -1 to 1 (1 = perfect agreement, 0 = chance agreement, ¡0 = less than chance). In our deep learning COVID-19 research, we will use Cohen’s Kappa to assess the agreement between our model’s predictions and the actual diagnoses of patients in order to accurately and reliably classify them as either “COVID-19 positive” or “COVID-19 negative”.

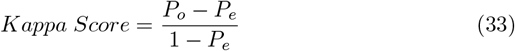 *P*_*o*_ represents the observed agreement, while *P*_*e*_ denotes for the expected agreement resulting from chance.
- **Area Under the Curve (AUC):** The area under the curve (AUC) is a fundamental notion that represents the integral of a curve concerning the x-axis within a specific range. The AUC-ROC is a performance measure used to assess the accuracy of binary classification models. The Receiver Operating Characteristic (ROC) Curve at this location illustrates the correlation between sensitivity and 1-specificity at various thresholds. The AUC-ROC varies between 0 and 1. A value of 0.5 signifies random guessing, while 1 implies perfect discrimination. Values between 0.5 and 1 denote performance that is superior to chance. Values below 0.5 indicate performance that is worse than random. ROC analysis evaluates the precision of medical diagnostic tests in differentiating between diseased and non-diseased conditions in clinical epidemiology [75].

## 4 Experimental Results and Discussion

To complete the entire experiment, we performed two experiments with two different types of data, one CT-scan image data and the other chest X-ray image data, to present our proposed model’s robustness. After training the model on the corresponding dataset, the trained model is saved in the Hierarchical Data Format version 5 format, which is mentioned as ‘.h5’ in the file name. This trained model can now be utilized for COVID-19 detection in the field of application. Since this format provides flexibility and support for data of any size, this saved model can detect COVID-19 by using CT scans and chest X-ray image data. This section discusses about the evaluation procedure carried out on the developed model to analyze its efficiency.

### 4.1 Performance on CT Scan dataset

We first experimented on the CT scan image dataset to evaluate our proposed model. To evaluate our model performance, we utilized a total of seven evaluation metrics that are Precision(Pre), Sensitivity(Sen), Specificity(Spe), Accuracy(ACC), F1-Score(F1-Sc), Cohen-Kappa Score(Co-Ka), and Area Under the Curve (AUC). In the model performance Table 3, we have used the short form of the evaluation metrics. The model’s accuracy represents the percentage of accurately classifying the images in the test dataset set. The model is trained with a sample of 1587 samples from the training dataset, validated with 397 samples, and tested with 497 sample images, which are split in proportion to the 60:20:20 ratio. The graph depicted in Figure 5 illustrates the progression of training accuracy, validation accuracy, training loss, and validation loss throughout each epoch of model training. At the beginning of training, during the first few epochs, both training and validation accuracies are poor, thus proving that the model does not train well at the beginning. Increasing the number of epochs quickly boosts the training accuracy to a high value, close to 1, indicating that it learns and fits the training data effectively. First, the validation accuracy is volatile, then it also grows and converges to a high value, one slightly lower than the model training accuracy.

**Fig 5.**
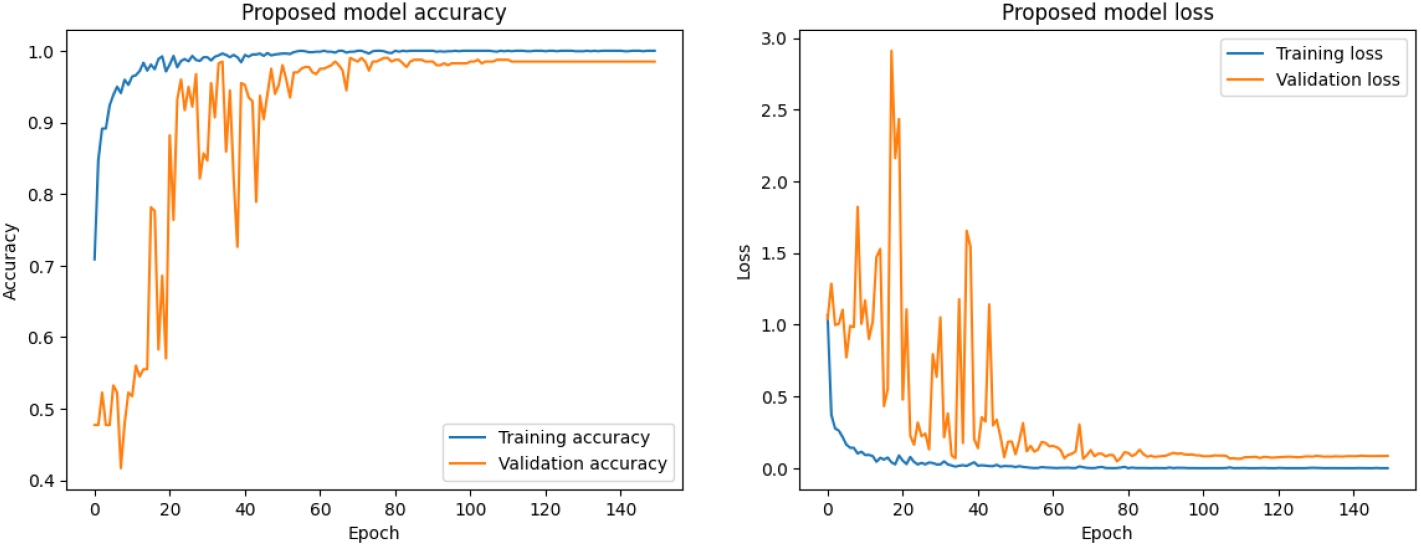
Proposed model accuracy and loss curve for CT-Scan Dataset

Probably due to either model initialization, learning rate, or noise in the data, it might be that the validation accuracy fluctuation occurs during the few epochs at the beginning. It tends towards the final stabilization near the training accuracy, indicating a good generalization to validation data with no significant model overfitting. The high accuracy of both the train data and the validation data proves to be quite good performance coverage. There is no overfitting against the training data when the training and validation accuracies have very little difference concerning the model; therefore, it is likely to perform well on unseen data.

A confusion matrix will help so much in knowing the performance of a classification model, contrasting true labels vs. predicted labels. In Figure 6, the Y-axis shows the true categories of the data. These categories are “COVID” and “non-COVID”, while the X-axis represents the categories predicted by the model. Top left, True Positives: 252 instances where the model correctly predicted “COVID”. Top-right: 1 instance of times where it has mispredicted “COVID” when it was actually “non-COVID”. Bottom left: 1 for the number of times the model predicted “non-COVID” where it is “COVID”.

**Fig 6.**
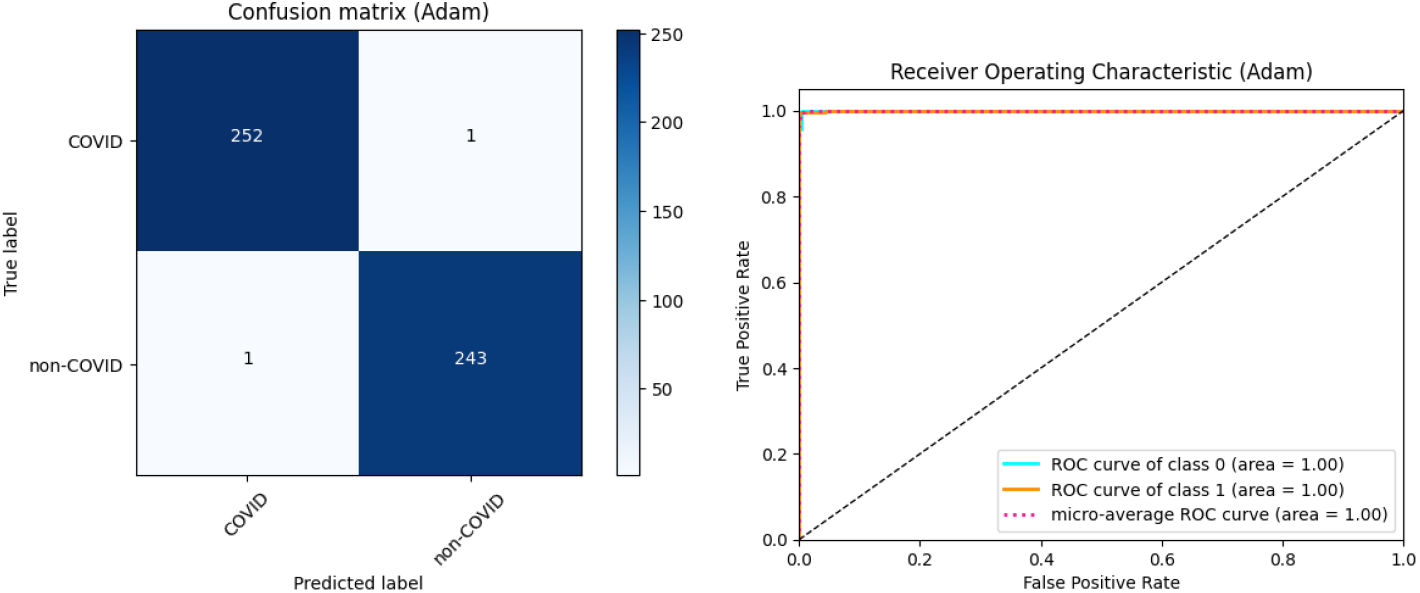
Proposed model confusion matrix and ROC curve for CT Scan Dataset

Bottom-right: True Negatives 243 cases where the model rightly predicted “non-COVID”. The performance of the model in classifying COVID-19 and non-COVID cases is very good, with very high accuracy, precision, recall, and high specificity. This means very minimal false positives and zero false negatives, which proves the model is very reliable for identifying true cases of COVID-19 without missing any. High values in the true positive and true negative cells and near-zero values in the false positive and false negative cells demonstrate that this is a very strong model.

Figure 6 represents the Receiver Operating Characteristic Curve (ROC) for a Classification Model. In this Figure, the X-axis represents the false positive rate, that is, the proportion of actual negatives that are misclassified as positives; the Y-axis represents the true positive rate, that is, the proportion of actual positives correctly classified as positives. The ROCs of both classes, 0 (cyan) and class 1 (orange), have an AUC of 1.00, which is a perfect classification with no false positives or false negatives. Also, the micro-average ROC curve is pink dashed with an AUC of 1.00, suggesting that the performance is uniform across classes. The random classifier is represented by the black dashed diagonal, where TPR=FPR and its AUC is 0.5. By contrast, ROC curves for the model under evaluation are well above this line, reflecting a high degree of discrimination. The ideal AUC scores with perfect separations between the classes show very good capability of the model in class discrimination, though such results may call for further investigation to check for overfitting in real-world applications. The combination of these curves presents a clear depiction of the model’s ability to achieve flawless sensitivity and specificity.

Table 2 shows the classification reports of the proposed model with three different optimizers. Among these optimizers, the Adam optimizer shows comparatively better results than the other optimizers. Table 3 shows performance metrics for various deep learning models, which have been optimized using different optimization algorithms on several evaluation criteria. The proposed model achieved the highest performance in all optimization metrics, mainly in Adam optimizers. It is followed that the metrics of precisions, sensitivity, specificity, accuracy, F1-Score, Cohen’s kappa, and AUC are highest for the proposed model are 99.60%, 99.59%, 99.60%, 99.60%, 99.60%, 99.16%, and 99.60% respectively. Among the three optimizers, RMSProp and SGD optimizers achieved the same sensitivity value of 99.55%. From Figure 7, we can easily compare the results for different optimizers individually for the CT scan dataset. This overall performance shows the proposed model to be of superior capability in predicting accurately and reliably on the desired outcomes.

**Table 2.**
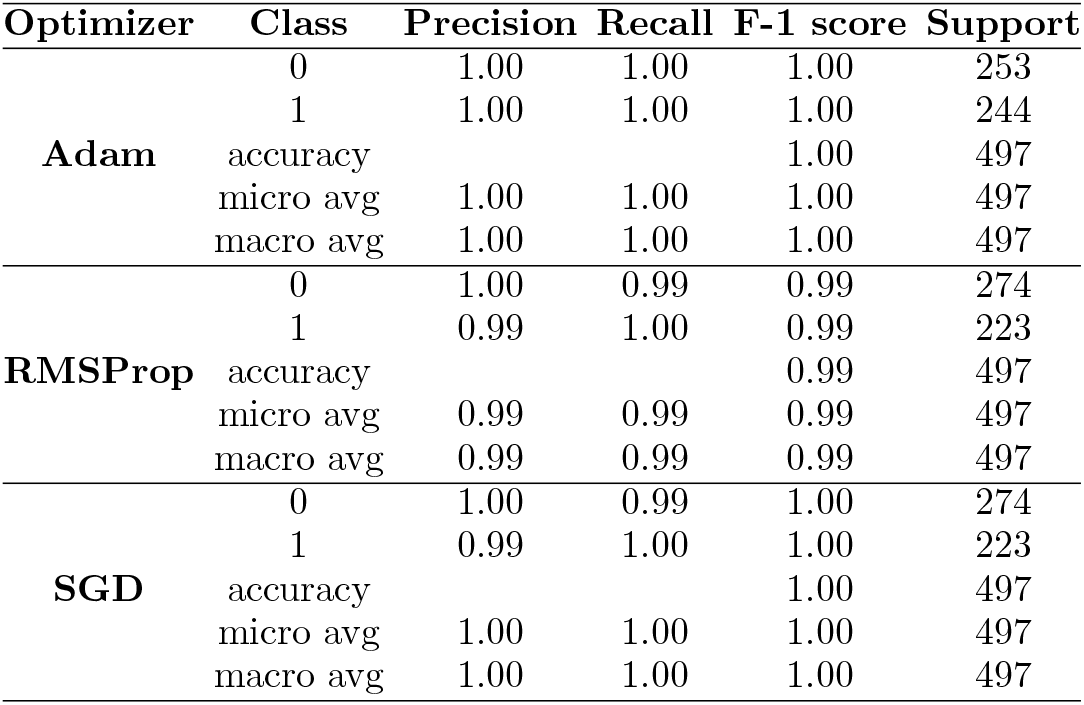
Classification Reports on CT-Scan Dataset.

**Table 3.** Proposed Model Classification Results for CT-Scan Image dataset.

| Model Name | Opt | Pre (%) | Sen (%) | Spe (%) | Acc (%) | F1-Sc (%) | Co-Ka (%) | AUC |
| --- | --- | --- | --- | --- | --- | --- | --- | --- |
| AlexNet | Adam | 91.22 | 94.17 | 89.05 | 91.35 | 91.31 | 82.64 | 91.61 |
|  | RMSProp | 82.31 | 90.58 | 74.09 | 81.49 | 81.49 | 63.35 | 82.34 |
|  | SGD | 91.20 | 97.76 | 85.40 | 90.95 | 90.94 | 81.96 | 91.58 |
| VGG16 | Adam | 92.21 | 92.83 | 91.97 | 92.35 | 92.29 | 84.58 | 92.40 |
|  | RMSProp | 89.78 | 91.48 | 88.69 | 89.94 | 89.99 | 79.77 | 90.08 |
|  | SGD | 83.51 | 82.96 | 84.31 | 83.70 | 83.56 | 67.13 | 83.63 |
| ResNet-50 | Adam | 98.37 | 98.21 | 98.54 | 98.39 | 98.37 | 96.75 | 98.37 |
|  | RMSProp | 98.56 | 98.65 | 98.54 | 98.59 | 98.58 | 97.15 | 98.60 |
|  | SGD | 98.12 | 98.65 | 97.81 | 98.19 | 98.17 | 96.34 | 98.23 |
| Xception | Adam | 93.24 | 96.41 | 90.88 | 93.36 | 93.33 | 86.67 | 93.64 |
|  | RMSProp | 79.63 | 84.31 | 75.55 | 79.48 | 79.45 | 59.06 | 79.93 |
|  | SGD | 76.37 | 78.48 | 74.82 | 76.46 | 76.37 | 52.83 | 76.65 |
| MobileNet | Adam | 95.66 | 96.41 | 95.26 | 95.77 | 95.74 | 91.48 | 95.83 |
|  | RMSProp | 95.09 | 95.07 | 95.26 | 95.17 | 95.12 | 90.25 | 95.16 |
|  | SGD | 93.46 | 93.27 | 93.80 | 93.56 | 93.50 | 87.00 | 93.53 |
| InceptionV3 | Adam | 98.37 | 98.55 | 98.27 | 98.40 | 98.39 | 97.78 | 98.41 |
|  | RMSProp | 98.93 | 99.55 | 98.54 | 98.99 | 98.98 | 97.97 | 99.05 |
|  | SGD | 99.05 | 98.21 | 99.64 | 98.99 | 98.98 | 97.96 | 98.92 |
| DenseNet121 | Adam | 93.83 | 94.62 | 93.43 | 93.96 | 93.91 | 87.83 | 94.02 |
|  | RMSProp | 90.46 | 94.17 | 87.59 | 90.54 | 90.51 | 81.05 | 90.88 |
|  | SGD | 83.69 | 87.44 | 80.66 | 83.70 | 83.66 | 67.40 | 84.05 |
| EfficientNetB0 | Adam | 92.10 | 83.41 | 97.45 | 91.15 | 90.91 | 81.88 | 90.43 |
|  | RMSProp | 91.27 | 90.13 | 92.34 | 91.35 | 91.25 | 82.50 | 91.24 |
|  | SGD | 97.05 | 99.10 | 95.62 | 97.18 | 97.16 | 94.33 | 97.36 |
| **Proposed** | Adam | **99.60** | **99.59** | **99.60** | **99.60** | **99.60** | **99.16** | **99.60** |
|  | RMSProp | 99.37 | 99.55 | 99.27 | 99.40 | 99.39 | 98.78 | 99.41 |
|  | SGD | 98.93 | 99.55 | 98.54 | 98.99 | 98.98 | 97.97 | 99.05 |

**Fig 7.**
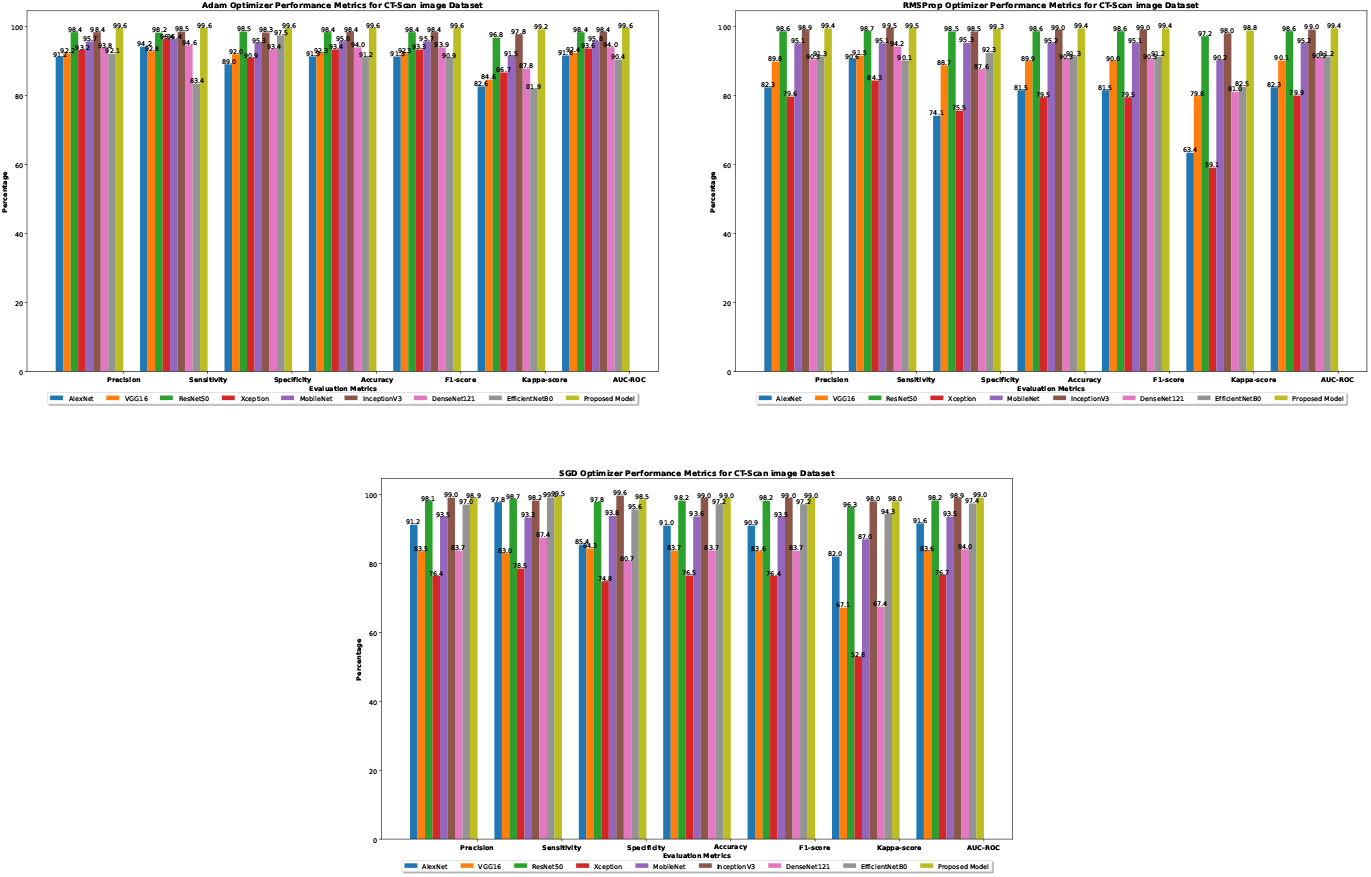
Comparison Bar Chart for different Evaluation Metrics for CT Scan Image dataset results with corresponding Models

### 4.2 Performance on X-ray dataset

For the second study, we have utilized the X-ray image dataset to classify COVID-19. In this dataset, we have included three class images, including COVID-19, pneumonia, and normal. We have used 12,576 images for the entire dataset, of which 80 percent have been used for training. Also, 20 percent of images from training data were used for the model validation. The remaining 20 percent have been utilized for the model testing, which was new for the model.

Figure 8 describes the performance of the proposed model on the chest X-ray dataset at 100 epochs, where one is about accuracy and the other one is about loss during training and validation. The training accuracy curve shows that the training accuracy starts low but increases rapidly to a value close to 1.0 (or 100%) by epoch 20, thus suggesting that the model can learn the training data very well. The validation accuracy curve is more volatile, especially at early epochs; it converged at about epoch 100. The validation accuracy also increased by nearly 1.0, indicating good generalization performance. These big swings in validation accuracy in the first epochs likely result from model adjustments, changes in learning rate, or data variability. With time, these fluctuations decrease. One can notice that the training loss begins high and reduces very fast initially, leveling near 0 at epoch 20 in the training loss curve. This means the model is efficiently reducing errors in the training data. The validation loss curve also illustrates quite a significant fluctuation, more so in the early epochs. Still, these fluctuations decrease over time, and the loss finally rests at a low value around epoch 100. The high and fluctuating validation loss in the initial epochs could result from overfitting or the model being fit to the distribution of validation data.

**Fig 8.**
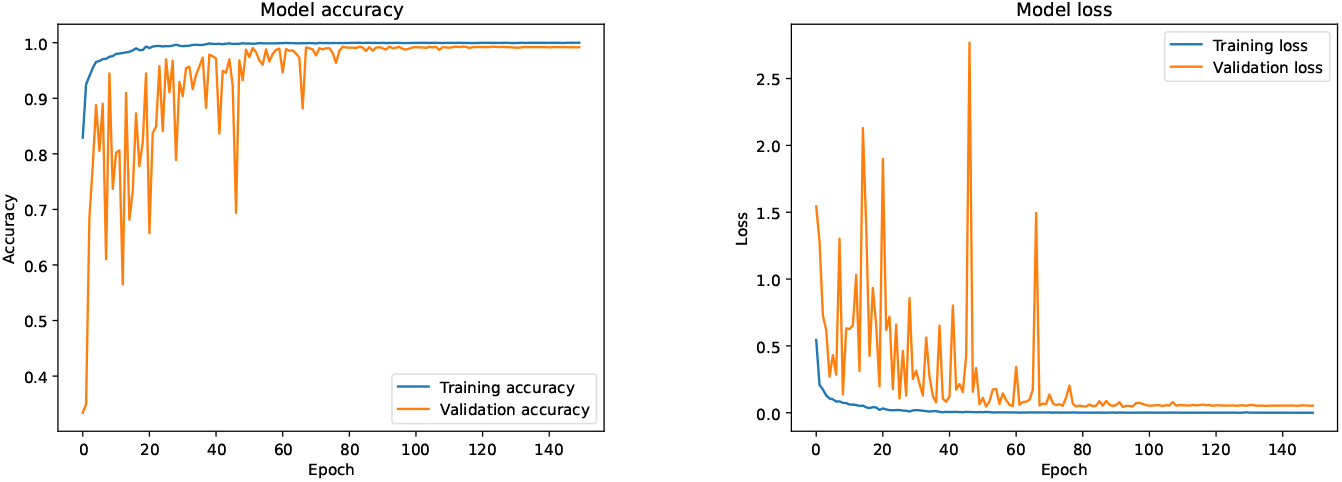
Proposed model accuracy and loss curve for X-ray Image Dataset

The fluctuations decrease, finally stabilizing at a low loss, so it must improve model performance and generalize better. The accuracy and loss plots indicate that the model converges with increasing training. The stabilization of training and validation accuracy close to 1.0, with loss reduction close to 0, may be viewed as proof of the model learning effectively and generalizing well. Instability of validation metrics, as manifested through accuracy and loss at the beginning, is quite a common phenomenon, denoting adjustment of parameters by the model. The eventual stabilization could, therefore, be taken to imply that these adjustments are practical in nature. High final accuracy and low final loss in both the training and validation sets imply that the model does not overfit and generalizes well to new data. These figures demonstrate that the model trains well, learns the data properly, and generalizes well, which is evidenced by the fact that training and validation metrics converge.

In Figure 9, the left figure represents the confusion matrix, which summarizes how well the multiclass classification model did on the three classes: COVID-19, pneumonia, and normal. It also perfectly represents how well the model differentiates between these three classes based on actual labels versus predicted labels. From this figure, 842, 824, and 821 were instances where models correctly predicted Covid-19, pneumonia, and normal, respectively. High true positive rates of the proposed model suggest that most instances of COVID-19, pneumonia, and normal cases are classified correctly. A high number of true positives in each category evidences this. The model misclassifies very few, hence highly reliable. Specifically, only 2 case of COVID-19 is miss-classified as Pneumonia and 1 as Normal; only 3 of Pneumonia are misclassified as COVID-19 and 5 as Normal; and only 17 of Normal are misclassified as COVID-19 and 1 as Pneumonia. This low miss-classification number within all categories proves the strength and efficiency of the model in differentiating between different respiratory conditions. This is strongly supported by the confusion matrix, which shows that the classification model is quite effective and reliable. It provides excellent performances in each class, COVID-19, pneumonia, and normal, with great true positive rates and negligible cases of false positives and false negatives. This means that this model is one such measure through assessment metrics for model performance. This also proves that the model is very apt at making accurate diagnoses of these conditions from only the input data itself.

**Fig 9.**
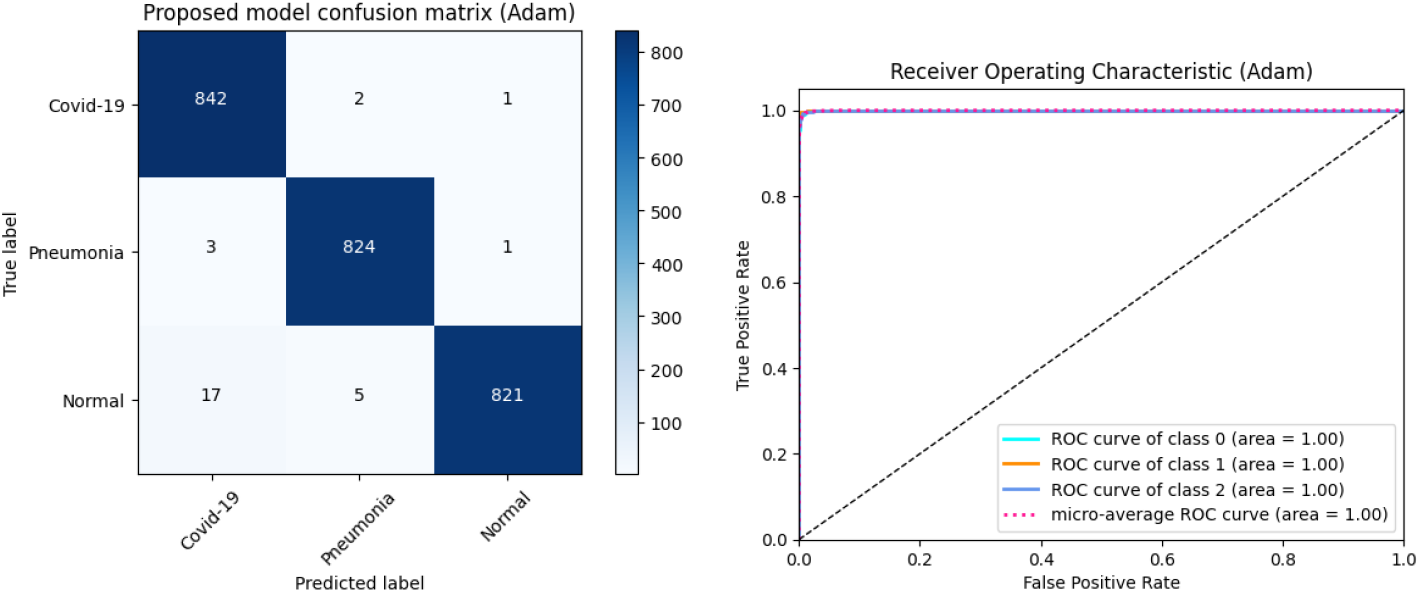
Proposed model confusion matrix for X-ray Image Dataset

In Figure 9, the figure of the Receiver Operating Characteristic curve is a graph showing the diagnostic ability of the classification model to make distinctions among three different classes: COVID-19, Pneumonia, and Normal. The ROC curve indicates the tradeoff between the TPR and the FPR at various threshold settings. The dashed diagonal line from the point (0,0) to the point (1,1) represents a random classifier. Any ROC curve that runs above this line suggests a model performing greater than a random chance. The ROC curve is plotted for three classes altogether representing class 0, class 1, and class 2 belonging to Covid-19, Pneumonia, and Normal cases, respectively. Cyan, orange, and blue curves represent the ROC curves corresponding to each class. These curves plot the relationship between TPR and FPR for various thresholds. The micro-average ROC curve, shown in pink dashed line, aggregates the performance across all classes. This is useful in the case of multi-class classification tasks. For each ROC curve, the AUC, or Area Under the Curve, is given in the legend. The AUC gives a single number summarizing the performance of the model. In the following plot, the AUC for all three classes is 1.00, which means the model is perfectly separating the classes. The three classes are perfectly classified with AUCs of 1.00, and there is complete separation between true positives and false positives, where no class cases are superimposed in their distribution. Actually, all three classes’ ROCs lie almost completely on the top-left corner of the plot, which is the ideal position a classifier can achieve. The micro-average ROC curve also illustrates an AUC of 1.00, reflecting the aggregate performance across all classes and confirming the overall strong performance of the model on the task of multi-class classification. From the receiver operating characteristic curves, it is evident that this model is quite promising for modularity in sensitivity-sensitivity trade-offs when predicting these conditions from a given dataset. This is a good pointer to the model’s reliability and that it can be deployed in the real world for diagnostic purposes since it can confidently distinguish the three respiratory conditions.

Table 4 shows the classification reports of the proposed model with three different optimizers for the X-ray dataset. In chest X-ray classification reports, among these optimizers, the Adam optimizer shows comparatively better results than the other optimizers. Table 5 shows the classification results prepared for various models over an X-ray image dataset. It provides an overall comparison of the performance of each model that applies three variously used optimization algorithms — Adam, RMSProp, and SGD — against multiple evaluation metrics. The table shows the full comparison of multiple models or optimizers and how each combination performs over an X-ray image dataset. Above, it can be noticed that the proposed model is that for which the Adam optimizer has overall performance with precision 98.97%, sensitivity 99.41%, specificity 99.40%, accuracy 99.40%, F1-Score 98.97%, Cohen’s Kappa 98.45%, and AUC 99.22%. Also, the RMSProp optimizer turned out to have very high performance in terms of specificity, and other metrics a bit lower than Adam on some metrics. The SGD optimizer also showed good performance, which was very close to Adam’s results. From Figure 10, we can easily compare the results for different optimizers individually for X-ray image dataset performance. The proposed model always performs best compared to all metrics and optimization algorithms, especially based on Adam. Generally, Adam provides the best performance across most models, showing higher precision, sensitivity, specificity, accuracy, F1-Score, Cohen’s Kappa, and AUC. Once again, RMSProp and SGD perform very competitively in a few models. In a few models, Adam fails to surpass SGD’s performance, where ResNet-50, InceptionV3, and DenseNet121 are found to report very good performance with each optimizer, which manifests and prove prepotency, efficiency, and robustness for handling and dealing with the dataset. With very near-to-perfect performance metrics recorded, it can now be confirmed that the proposed model is very effective and reliable for classifying the X-ray images.

**Table 4.** Classification Reports on Chest X-ray Dataset.

| Optimizer | Class | Precision | Recall | F-1 score | Support |
| --- | --- | --- | --- | --- | --- |
| **Adam** | 0 | 0.99 | 0.99 | 0.99 | 845 |
|  | 1 | 1.00 | 0.99 | 0.99 | 828 |
|  | 2 | 0.99 | 0.99 | 0.99 | 843 |
|  | accuracy |  |  | 0.99 | 2516 |
|  | micro avg | 0.99 | 0.99 | 0.99 | 2516 |
|  | macro avg | 0.99 | 0.99 | 0.99 | 2516 |
| **RMSProp** | 0 | 1.00 | 1.00 | 1.00 | 845 |
|  | 1 | 0.99 | 1.00 | 0.99 | 828 |
|  | 2 | 1.00 | 0.99 | 1.00 | 843 |
|  | accuracy |  |  | 1.00 | 2516 |
|  | micro avg | 1.00 | 1.00 | 1.00 | 2516 |
|  | macro avg | 1.00 | 1.00 | 1.00 | 2516 |
| **SGD** | 0 | 0.98 | 0.99 | 0.99 | 845 |
|  | 1 | 0.99 | 0.99 | 0.99 | 828 |
|  | 2 | 0.99 | 0.98 | 0.98 | 843 |
|  | accuracy |  |  | 0.99 | 2516 |
|  | micro avg | 0.99 | 0.99 | 0.99 | 2516 |
|  | macro avg | 0.99 | 0.99 | 0.99 | 2516 |

**Table 5.** Proposed Model Classification Results for X-ray Image dataset.

| Model Name | Opt | Pre (%) | Sen (%) | Spe (%) | Acc (%) | F1-Sc (%) | Co-Ka (%) | AUC |
| --- | --- | --- | --- | --- | --- | --- | --- | --- |
| AlexNet | Adam | 89.12 | 85.33 | 93.48 | 90.74 | 88.91 | 83.18 | 91.57 |
|  | RMSProp | 90.33 | 91.60 | 91.26 | 91.38 | 90.11 | 85.15 | 92.56 |
|  | SGD | 90.06 | 90.41 | 91.68 | 91.26 | 89.93 | 84.80 | 92.38 |
| VGG16 | Adam | 94.57 | 93.14 | 96.29 | 95.23 | 94.55 | 91.77 | 95.88 |
|  | RMSProp | 93.14 | 90.77 | 95.45 | 93.88 | 93.15 | 89.69 | 94.83 |
|  | SGD | 82.68 | 83.20 | 90.31 | 87.92 | 82.38 | 73.28 | 86.62 |
| ResNet-50 | Adam | 98.17 | 98.05 | 98.70 | 98.48 | 98.17 | 97.75 | 98.37 |
|  | RMSProp | 97.81 | 97.93 | 97.40 | 97.24 | 97.81 | 97.21 | 97.11 |
|  | SGD | 98.09 | 97.58 | 98.70 | 98.32 | 98.09 | 97.63 | 98.31 |
| Xception | Adam | 93.92 | 91.60 | 96.47 | 94.83 | 93.91 | 90.82 | 95.40 |
|  | RMSProp | 93.23 | 90.53 | 96.23 | 94.32 | 93.20 | 89.75 | 94.86 |
|  | SGD | 93.70 | 90.65 | 96.59 | 94.59 | 93.67 | 90.46 | 95.22 |
| MobileNet | Adam | 95.11 | 94.32 | 96.89 | 96.03 | 95.12 | 92.67 | 96.33 |
|  | RMSProp | 95.32 | 92.19 | 98.08 | 96.10 | 95.29 | 92.91 | 96.45 |
|  | SGD | 95.33 | 91.60 | 98.20 | 95.99 | 95.22 | 92.79 | 96.39 |
| InceptionV3 | Adam | 98.57 | 98.82 | 99.04 | 98.97 | 98.57 | 97.85 | 98.93 |
|  | RMSProp | 98.50 | 99.17 | 98.86 | 98.97 | 98.49 | 97.73 | 98.87 |
|  | SGD | 98.69 | 98.06 | 98.70 | 98.68 | 98.05 | 97.08 | 98.54 |
| DenseNet121 | Adam | 95.27 | 93.73 | 97.37 | 96.14 | 95.22 | 92.79 | 96.38 |
|  | RMSProp | 94.98 | 92.19 | 97.43 | 95.67 | 94.94 | 92.37 | 96.18 |
|  | SGD | 94.65 | 91.95 | 97.37 | 95.55 | 94.59 | 91.83 | 95.91 |
| EfficientNetB0 | Adam | 83.10 | 80.00 | 93.72 | 89.11 | 81.26 | 72.54 | 86.29 |
|  | RMSProp | 88.56 | 80.00 | 94.91 | 89.90 | 88.21 | 82.29 | 91.13 |
|  | SGD | 82.07 | 77.75 | 90.31 | 86.09 | 80.69 | 70.71 | 85.33 |
| **Proposed** | Adam | **98.97** | **99.41** | **99.40** | **99.40** | **98.97** | **98.45** | **99.22** |
|  | RMSProp | 98.89 | 99.05 | 99.34 | 99.24 | 98.89 | 98.33 | 99.16 |
|  | SGD | 98.66 | 98.46 | 99.28 | 99.01 | 98.65 | 97.97 | 98.98 |

**Fig 10.**
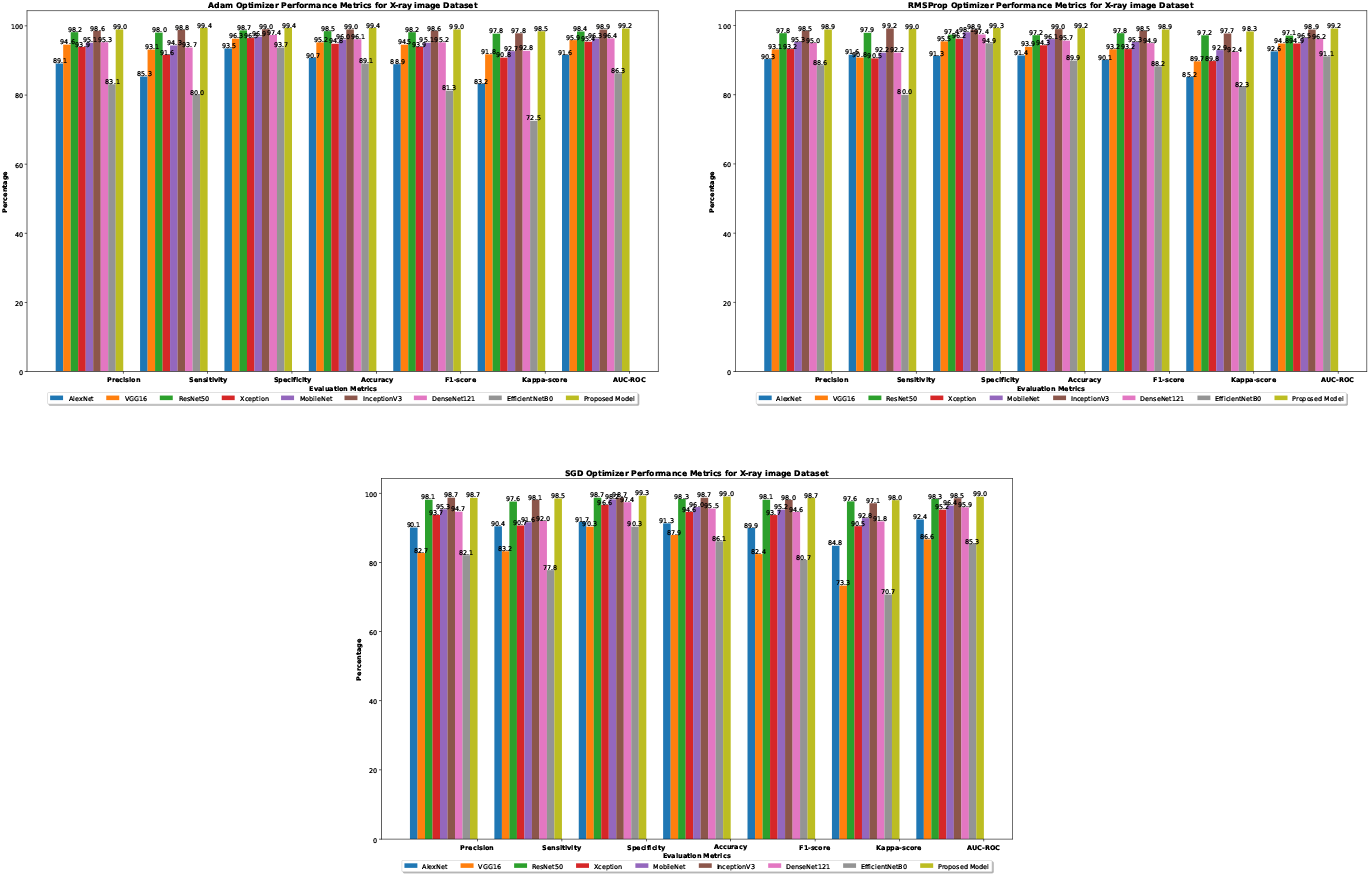
Comparison bar chart for different optimizers for chest x-ray dataset results with corresponding models

### 4.3 Proposed model with explainable AI

Grad-CAM enhances interpretability by highlighting the most important features or areas in a model’s decision-making process; therefore, this enhances better transparency and reliability of a model’s decisions. Figure 11 and Figure 12 show the application of Grad-CAM in CT scan images and chest X-ray images, respectively. The area is highlighted for consideration during diagnosis. Original CT scans are shown on the left, depicting cross-sectional views of the human body. Regions can be underlined that are affected by abnormalities such as lesions or infections. Grad-CAM heatmaps on the right overlay these images with color gradients that show areas of interest to be most relevant according to the deep learning model for its predictions. Warmer indicates yellow to red indicates regions of high importance, while cooler colors represent areas of lesser importance. First set: In this, the model underlined great areas within the lungs, reinforcing that the attention of the model is on possible abnormalities related to the disease, such as fibrosis or tumors. Second set: This unmasks another focus where the underlined heatmap does in other anatomical areas, probably referring to other pathological features. Figure 12 shows chest X-ray images; from the heatmap derived from the first pair of images, it would appear that this model focuses on the upper regions of both lungs, which might point toward an abnormality that could be fibrosis or consolidation. The second pair of images shows a more focused area of interest in the middle zone of the lung, which may indicate a suspicious lesion or infection. The third pair focuses strongly on the model in the right lung with red and yellow colors, which stand for a conspicuous feature that might be a pathological one. This visualization underlines Grad-CAM’s utility in enhancing the interpretability of deep learning models: giving medical practitioners insight into how the model interprets the complex diagnostic data and ensuring that focus is clinically relevant and aligned with human expertise.

**Fig 11.**
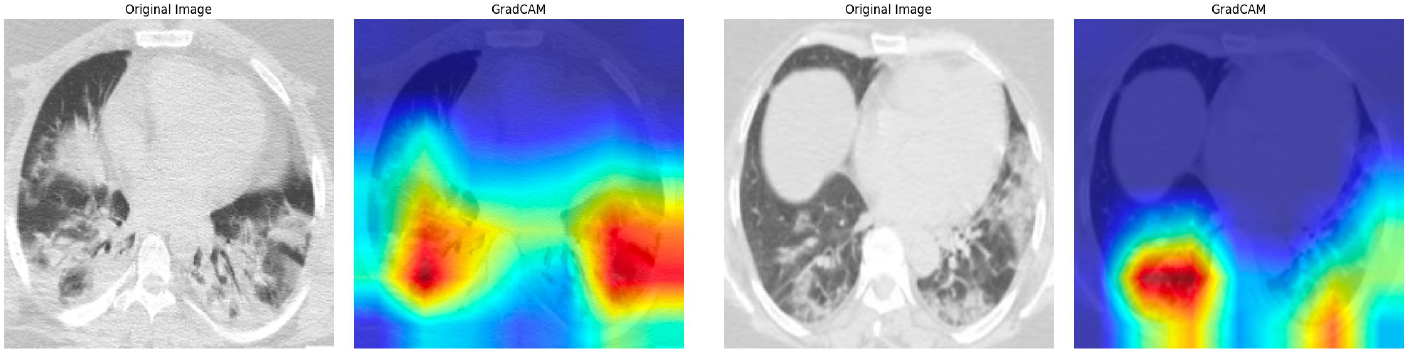
GradCAM-based visualization for the proposed model for CT-Scan image

**Fig 12.**
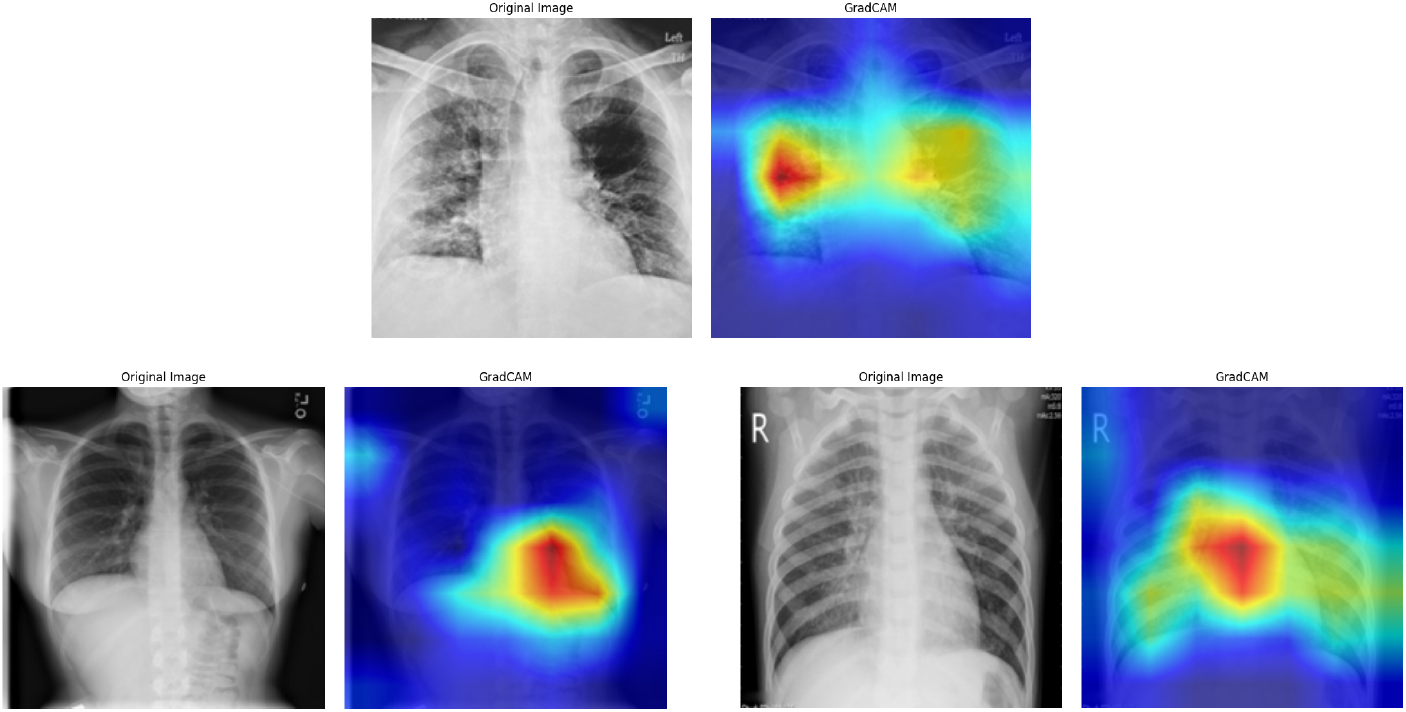
GradCAM-based visualization for the proposed model for X-ray image

LIME helps interpret the model by slightly changing the input data and observing how the predictions change, highlighting the areas that contribute most to the model’s decision. Figures use Local Interpretable Model-agnostic Explanations (LIME) to explain the model’s prediction on CT scans and X-ray images. The far left image in Figure 13 is the original CT Scan with the prediction from the model. A prediction value of 1.00 indicates a very high confidence in the prediction—COVID-19 in this case. Next image with all superpixels: This CT scan is segmented into superpixels, contiguous regions of similar pixel intensities. LIME uses such superpixels to perturb the image and see how much the model cares about changes in these regions. The positive superpixels image emphasizes the superpixels that positively contribute to the model’s prediction. These are very fundamental areas on which the model builds a lot of confidence in predicting the class. Here, different colors indicate the degree of importance; warmer colors, like red, stand for large positive contributions. The heatmap shows a gradient of contributions by several superpixels. There is a color scale that signals the strength of their contribution. The scale ranges from negative to positive contributions. Here, warmer colors show the most important positive contributions, while cooler colors like blue are for negative or unimportant contributions. This is how the X-ray image could be explained in the same way. Positive Superpixel images indicate areas that most positively influence model predictions. For both CT scans and X-rays, regions highlighted in warm colors, like red, are important for a model’s decision. The Heatmap provides an exact view of how each superpixel contributes to the prediction, with the color intensity reflecting the strength of contribution. In this case, on the CT scan image, superpixels surrounding the lungs and specific patterns within the lungs show up as strong contributors to the model’s prediction. The highlighted regions in the Positive Superpixels and Heatmap images mark areas with possible abnormalities the model has engrafted to the predicted class. In Figure 14, in the case of the X-ray image, just like in the CT scan, superpixels within the lung regions are highly important. Indeed, the Positive Superpixels and Heatmap images reveal this: the model focused on patterns or anomalies within the lungs characteristic of the predicted class. LIME aids in building trust in the model’s predictions by providing a visual explanation of which parts of the image influence the decision. This could be especially helpful for medical practitioners to check if the model concentrates on clinically relevant areas.

**Fig 13.**
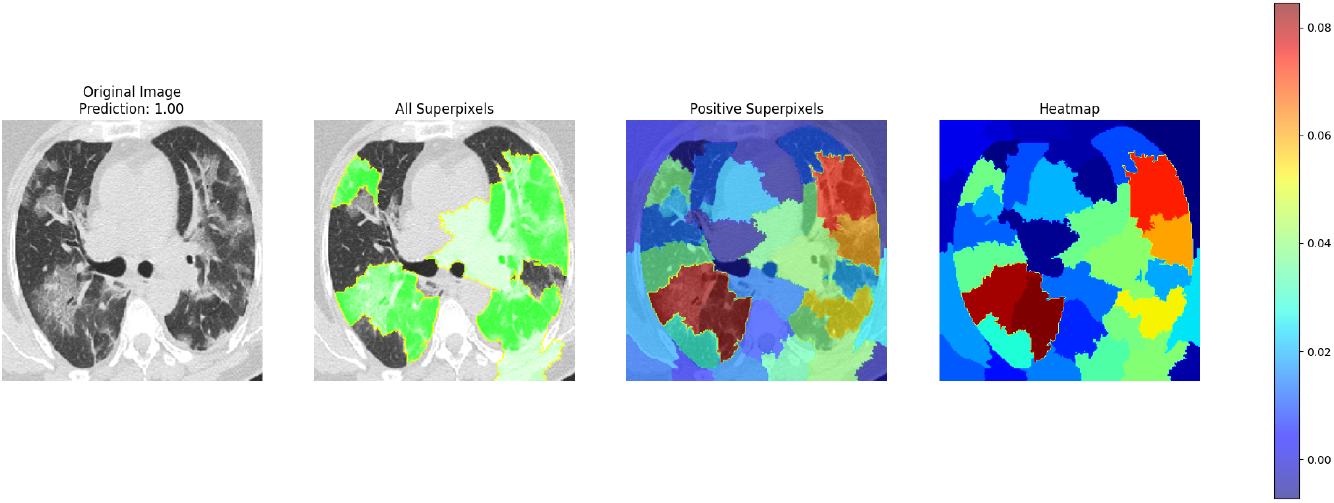
LIME-based visualization for the proposed model for CT scan image

**Fig 14.**
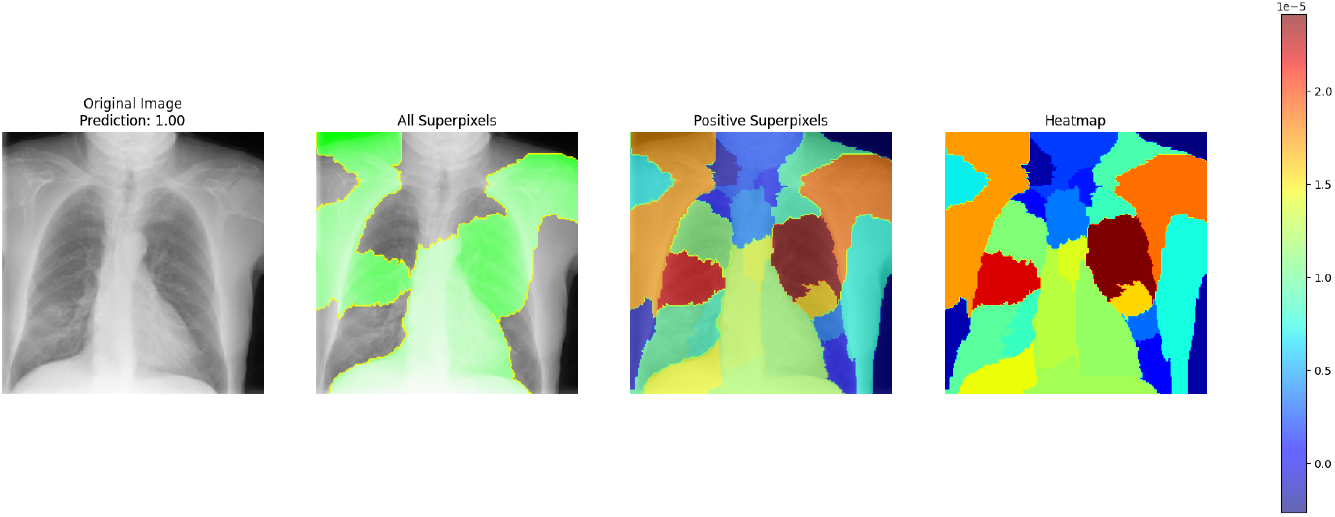
LIME-based visualization for the proposed model for X-ray image

Shapley’s Additive explanations are based on a game-theoretic approach that forces each feature to receive an important value for a particular prediction from the machine learning model. In Figure 15, here is one of the figures containing three images: one original CT scan image for a patient’s lungs, the other two being the SHAP values for two different classes, non-COVID and COVID. The input image serves as a reference for the areas being analyzed for the presence of COVID-19 or non-COVID conditions. The middle picture presents the SHAP values overlay on top of the CT scan for the non-COVID class. In the picture, blue regions correspond to a negative SHAP value and thus contribute to the model prediction of the image as non-COVID. The red areas are those parts where SHAP values are positive, contributing less towards the non-COVID prediction. The color intensity corresponds to the magnitude of SHAP values, with darker shades meaning higher absolute values. The right image projects the SHAP values onto the CT scan for the COVID class. In the image, red regions indicate positive SHAP values, and they contribute to the model predicting the image as COVID.

**Fig 15.**
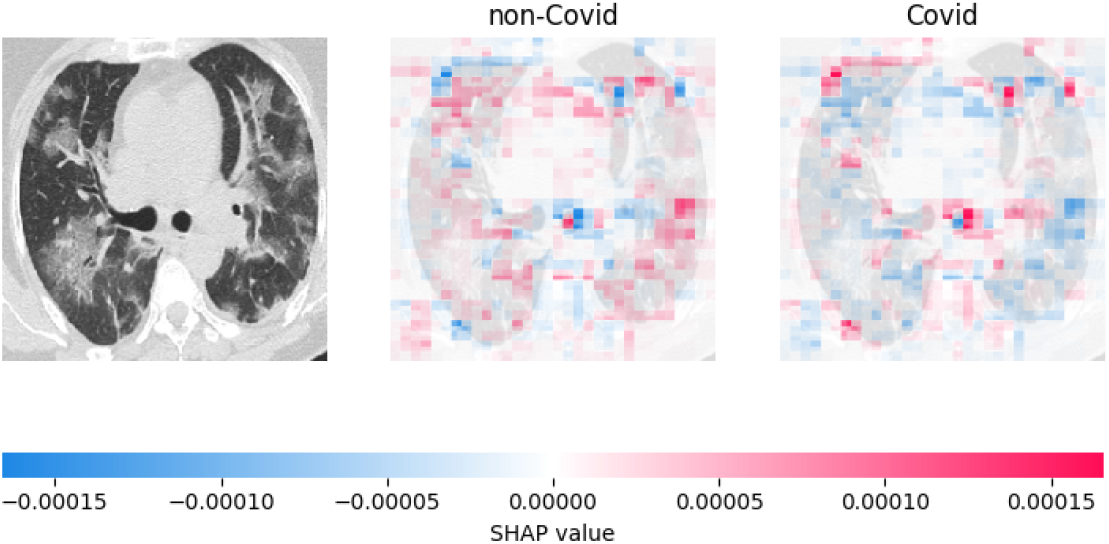
SHAP-based visualization for the proposed model for CT scan image

In contrast, the blue region has a negative SHAP value, contributing less to COVID prediction. The intensities of the colors represent the magnitude of the SHAP values; the darker shades indicate higher absolute values. The color bar gives a scale for interpreting the SHAP values. The farther away the SHAP value from zero, the stronger the contribution of that region to the prediction (-0.0002 or 0.0002). The middle and right images identify which areas of the CT scan are very important to have the model’s predictions for non-COVID and COVID, respectively. For non-COVID, areas marked in blue (negative SHAP values) play a very important role in influencing the prediction toward non-COVID. For COVID, areas colored red (positive SHAP values) have a strong influence on the COVID prediction. By comparing the SHAP value overlays, one can see how different parts of the CT scan impact the model’s predictions for non-COVID vs COVID. If a region is red in the COVID-SHAP image and blue in the non-COVID-SHAP image, the combination makes that a very strong COVID indicator. SHAP values explain how the model works by showing the areas most influential in the classification of the CT scan as either COVID or non-COVID. This may be useful in understanding the strengths and weaknesses of the model.

Figure 16 contains four images: an x-ray image and its corresponding SHAP values for three different classes. We can explain our model with the same information for the three classes’ data as we explained for the CT scan images. Here the middle three images help to indicate which parts of the x-ray are most important to the model predictions for COVID-19, Pneumonia, and normal, respectively. In the COVID-19, red areas show a very strong positive influence of SHAP values on the prediction towards COVID-19. For Pneumonia, areas in red have positive SHAP values that strongly influence the prediction of Pneumonia. It means for Normal, areas in red marked with positive SHAP values strongly influence the prediction towards Normal.

**Fig 16.**
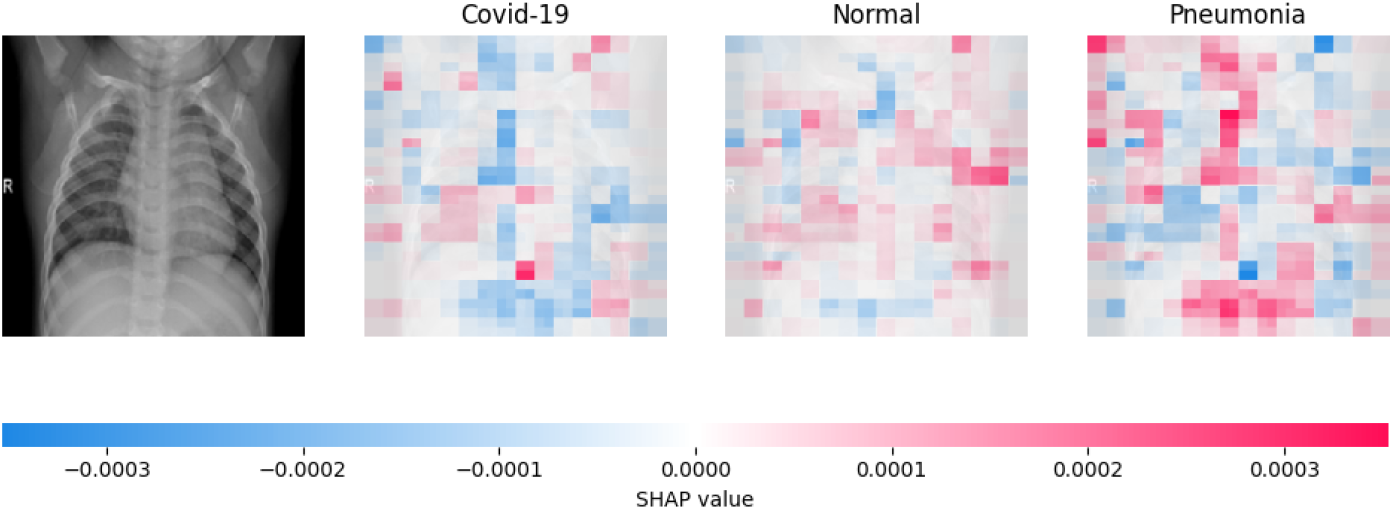
SHAP-based visualization for the proposed model for X-ray image

The SHAP figure properly visualizes how CT scan images in non-COVID and COVID conditions and X-rays in COVID-19, Pneumonia, and Normal conditions contribute different regions to classification. Areas of importance are highlighted with positive and negative SHAP values for explanation, clearly showing how the model made its predictions to interpret and trust its decisions in a clinical setting.

### 4.4 Model validation using User Interface (UI)

A user interface is developed according to the provided paradigm for the real-time validation of our suggested framework. The interface is developed using Gradio, an open-source Python package. This study utilized Python version 3.12.7 and Gradio version 2.9.2. Gradio facilitates the rapid creation of customizable web-based Graphical User Interfaces (GUIs) for deploying machine learning models or specific functions. This assists us in constructing a prototype of the deployed model to evaluate its performance in the user environment. The Gradio Interface fundamentally requires three parameters: the function defining the classification model, the input components, and the output components. Upon execution, a web page that includes the user interface is generated. The web page’s URL is shareable and can be disseminated publicly. Since this is publicly available, any individual with a browser can utilize it, provided the host device is operational. Figure 17 depicts the user interface wherein an image is uploaded and submitted for classification, yielding an output accompanied by the confidence level. This serves as user feedback for the model’s enhancement. To erase the current image, the ‘Clear’ button is activated. Various CT scans and chest X-ray images from different classes have been uploaded and tested in real-time, as illustrated in Figure 18. The model demonstrates 100% confidence in identifying CT scan pictures as COVID-19 or normal and chest X-ray images as COVID-19, pneumonia, or normal. Consequently, the suggested model exhibits significant performance in a real-time setting. The user can modify the uploaded image by cropping, rotating, or introducing noise to observe the variations in output.

**Fig 17.**
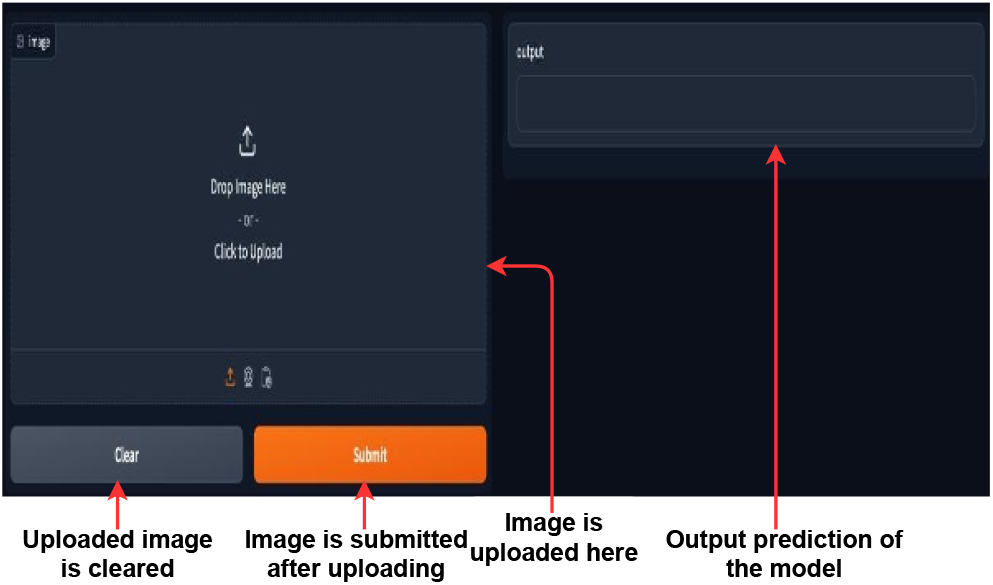
A Python Gradio user interface was designed to assess the proposed model’s real-time prediction capability.

**Fig 18.**
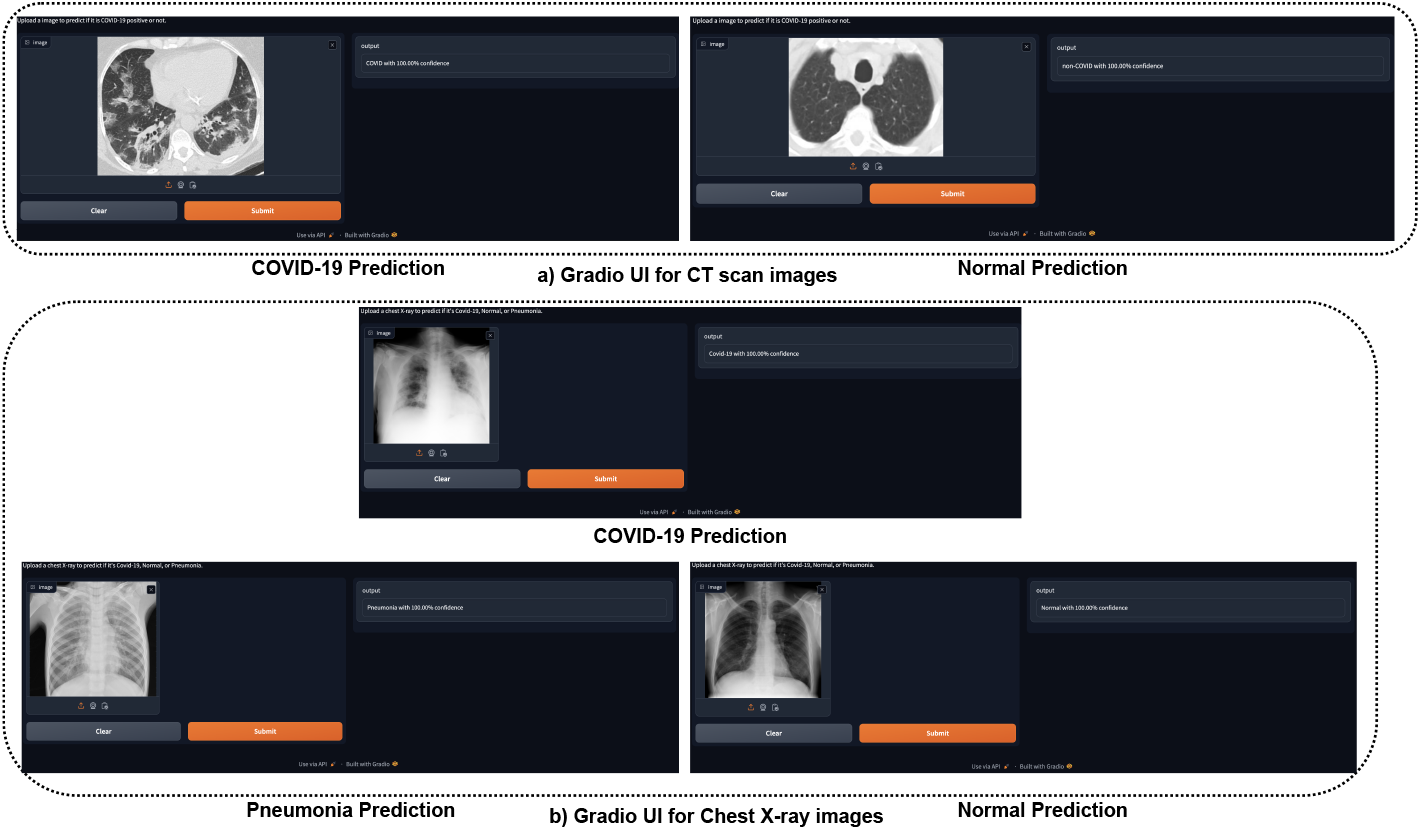
Gradio user interface (UI) for CT Scan and Chest X-ray image prediction.

## 5 Discussion

The findings we demonstrate illustrate the effectiveness of the RGFSAMNet model in precisely identifying and classifying COVID-19 utilizing datasets of CT scans and chest X-ray images. Exceptional accuracy rates were achieved, exceeding the results of earlier research employing pre-trained deep learning models such as AlexNet, VGG16, ResNet50, MobileNet, InceptionV3, DenseNet121, and EfficientNetB0. This indicates that our suggested RGFSAMNet model could be highly impactful in the field of medical imaging analysis, particularly in the identification and classification of COVID-19. The RGFSAMNet model’s exceptional accuracy is essential for the detection and classification of COVID-19. COVID-19 necessitates accurate diagnosis, akin to other disorders, to enhance patient outcomes and guide suitable treatment choices. The suggested model demonstrates enhanced performance by effectively extracting features from CT scans and chest X-ray images through a residual learning block incorporating feature fusion and an attention mechanism. Moreover, while evaluating various optimizers like Adam, RMSprop, and SGD, a minor discrepancy in performance was seen. Nonetheless, RGFSAMNet had high accuracy rates across all three optimizers, showing its robustness to the model.

Seven performance metrics—precision, accuracy, F1-score, sensitivity, specificity, Cohen-Kappa score, and area under the curve—have been estimated beyond accuracy. These measures provided a comprehensive assessment of the model’s performance to accurately identify negative cases, as well as the sensitivity and specificity of positive cases. The outcomes for all employed metrics consistently surpass those of traditional baseline models in the proposed RGFSAMNet model. This demonstrates its ability to accurately classify COVID-19 illness. Cohen’s Kappa was utilized to evaluate inter-rater agreement in assessing model performance. The elevated values of Cohen’s Kappa indicate a robust concordance between the model’s predictions and the actual labels. The model that was suggested generates consistent and accurate forecasts, so offering a good diagnosis. Table 6 offers a thorough accuracy and recall comparison of our model with current models. Table 6 shows that our model regularly outperforms others over several datasets, so indicating its higher efficiency than current approaches. The model’s effective generalization to unfamiliar datasets is evidenced by its robust performance on two relatively independent datasets for COVID-19 detection. This indicates that the model is crucial for efficiently processing new information, offering a dependable and versatile solution for various medical applications.

**Table 6.**
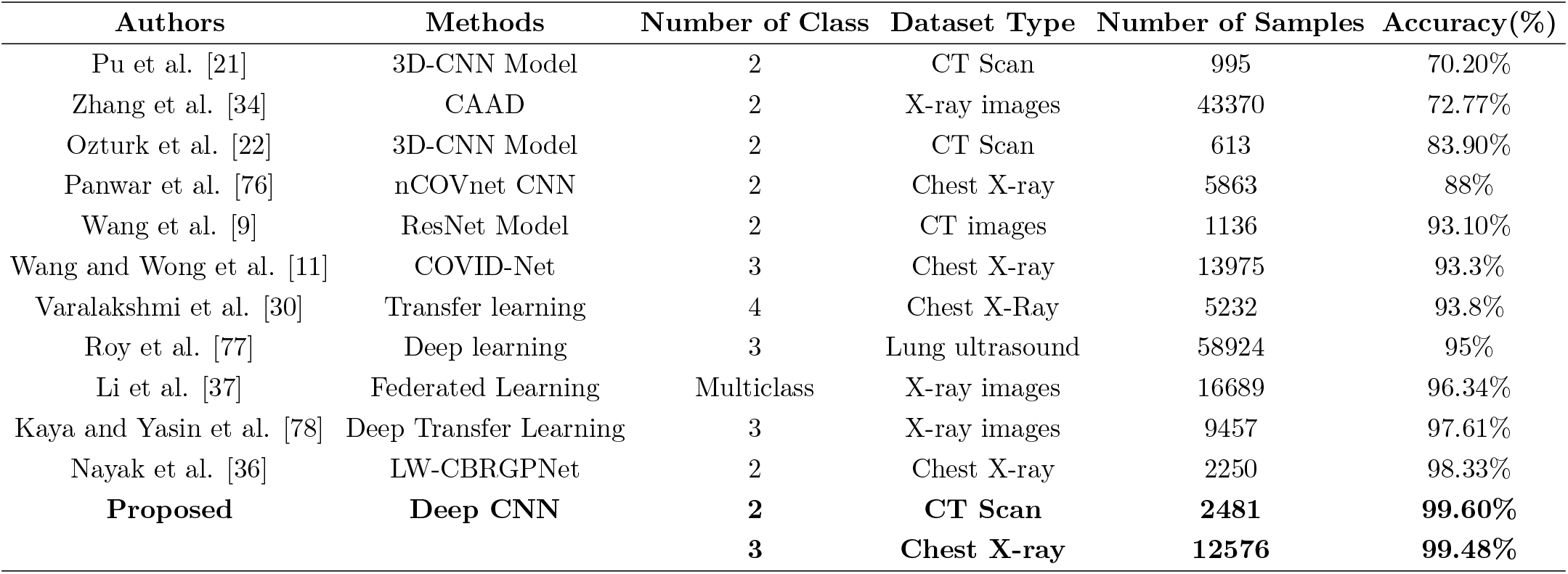
Comparison with previous existing state-art-of-the model performances.

We dedicated a significant portion of our work to enhancing the interpretability of the proposed model by incorporating diverse strategies from Explainable Artificial Intelligence (XAI). Grad-CAM, LIME, and SHAP provided insightful details regarding the precise areas of the CT scan and chest X-ray pictures that significantly influenced the model’s decision-making process. The outcome entails enhancing trust and facilitating in-depth scrutiny for doctors and researchers, leading to a more comprehensive comprehension of the model’s cognitive processes. The model’s remarkable efficiency in terms of training time is particularly notable. The training duration for this model was significantly decreased compared to the current models. Concerning this issue, the model is exceedingly practical for real-world applications. Extending the duration of training and producing unambiguous results can lead to a timely and efficient analysis of medical imaging data, thereby triggering a substantial transformation in healthcare and clinical decision-making. During the COVID-19 pandemic, the use of Grad-CAM, LIME, and SHAP techniques has brought attention to specific visual features of the disease, such as ground-glass opacities and consolidation patterns, as well as the degree of lung involvement found in CT scans and Chest X-rays. The data indicate that this model can precisely forecast COVID-19 infection in CT scans and Chest X-ray pictures. These approaches allow doctors to focus on critical locations while processing CT images, resulting in rapid and accurate diagnosis of COVID-19. Therefore, it is crucial to ensure the effective management and containment of diseases, mainly through prompt intervention and appropriate allocation of resources.

This study utilizes CT scans and chest X-ray images, employing the proposed RGFSAMNet model to thoroughly assess the challenges of detecting and classifying COVID-19. The model surpassed contemporary models in precision, accuracy, F1 score, sensitivity, specificity, Cohen-Kappa score, and area under the curve. By clarifying elements of the decision-making process, XAI techniques have improved the interpretability of the model. Therefore, the suggested model is quite likely to improve accuracy and efficiency in COVID-19 classification, so benefiting patients as well as doctors. More research on its validation over other datasets is advised to improve the dependability and applicability of the suggested model in a clinical environment.

## 6 Conclusion

Utilizing deep learning algorithms for categorization can expedite the identification of COVID-19 and other contagious illnesses. This paper presents an experimental evaluation of a deep residual network with global feature fusion and an attention mechanism-based image classification approach to enhance performance accuracy. This study aimed to develop a method for distinguishing COVID-19 infection using CT scans and chest X-ray images. Experimental results indicate that the improved RGFSAMNet model surpasses an individual model in terms of accuracy. This model achieves an accuracy of over 99.60% on lung CT scan datasets and an accuracy of 99.48% on chest X-ray image datasets. The results are promising because the proposed models will help experts diagnose quickly and accurately. The trained model is very cost-effective and user-friendly and can also be used in a clinic to diagnose patients. From experimental observation, it is clear that our proposed model and approach can have a massive impact on the spread of COVID-19 detection and also provide past screening.

## Data Availability

www.kaggle.com/datasets/plameneduardo/sarscov2-ctscan-dataset https://www.kaggle.com/datasets/tawsifurrahman/ covid19-radiography-database?fbclid= IwY2xjawEscmdleHRuA2FlbQIxMAABHbVnOUHP_IkOnkGs_ YT8gn3p9OcHUpEMovCm8SJRVSaX8SrK-uGOneT3fA_aem_lo2qRJKfp8k_ fvbCG5H-0w https://www.kaggle.com/datasets/prashant268/chest-xray-covid19-pneumonia?fbclid=IwY2[…]CL26aQKKwB3V1wg_kNTcnQlK7AM0DKpm5Q_aem_w5mc_8BA_60EUyQhv6JpRw

## Author Contributions

S.M.R.U.K.: Conceptualization, Methodology, Software, Writing original draft, review & editing, D.B.: Conceptualization, Methodology, Software, Data curation, Writing original draft, R.A.R.: Writing, review & editing, S.G.: Conceptualization, Methodology, Writing, review & editing and Supervision.

## Acknowledgments

The authors express their gratitude to the web portal organization and journalists who submitted COVID-19 images to the online resources.

